# Disentangling Maternal and Fetal Genetic Contributions to Preeclampsia

**DOI:** 10.64898/2025.12.03.25341531

**Authors:** Jaakko Leinonen, Jaakko Tyrmi, Tea Kaartokallio, A. Inkeri Lokki, FINNPEC study group, FINNGEN, Estonian Biobank Research Team, Triin Laisk, Anneli Pouta, Katja Kivinen, Eero Kajantie, Seppo Heinonen, Juha Kere, Tanja Saarela, Tiina Jääskeläinen, Hannele Laivuori

**Affiliations:** Center for Child, Adolescent and Maternal Health Research, Faculty of Medicine and Health Technology, Tampere University, Tampere, Finland; Center for Life Course Health Research, Faculty of Medicine, University of Oulu, Oulu, Finland; Department of Bacteriology and Immunology, University of Helsinki and Helsinki University Hospital, Helsinki, Finland; Estonian Genome Centre, Institute of Genomics, University of Tartu, Tartu, Estonia; Finnish Institute for Health and Welfare, Helsinki, Finland; Institute for Molecular Medicine Finland, Helsinki Institute of Life Science, University of Helsinki, Helsinki, Finland; PEDEGO Research Unit, Medical Research Center Oulu, Oulu University Hospital and University of Oulu, Oulu, Finland; Population Health Unit, Finnish Institute for Health and Welfare, Helsinki and Oulu, Finland; Children’s Hospital, University of Helsinki and Helsinki University Hospital, Helsinki, Finland; Department of Clinical and Molecular Medicine, Norwegian University of Health and Technology, Trondheim, Norway; Department of Obsterics and Gynaecology, University of Helsinki and Helsinki University Hospital, Helsinki, Finland; Department of Biosciences and Nutrition, Karolinska Institutet, Huddinge, Sweden; Folkhälsan Research Center and Stem Cells and Metabolism Research Program, University of Helsinki, Helsinki, Finland; Department of Clinical Genetics, Kuopio University Hospital, Wellbeing Services County of North Savo, Kuopio, Finland; Department of Food and Nutrition, University of Helsinki, Helsinki, Finland; Department of Obstetrics and Gynecology, Tampere University Hospital, The Wellbeing Services County of Pirkanmaa, Tampere, Finland

## Abstract

Hypertensive disorders of pregnancy are a leading cause of maternal and fetal morbidity and mortality, with preeclampsia (PE) affecting ∼5% of pregnancies worldwide. Both maternal and fetal genomes influence PE risk, but their relative contributions remain unclear. We conducted a meta-analysis of genome-wide association studies assessing maternal and fetal genetic effects on PE, including 401,597 maternal and 435,076 fetal samples. We identified 27 independent PE-associated variants, comprising 12 novel and 3 fetal-specific loci. Maternal and fetal genetic effects were independent, potentially reflecting opposing selective pressures on maternal versus fetal determinants of PE. Most heritable risk arose from maternal variants linked to blood pressure regulation, adiposity, and immune tolerance. Causality analyses supported roles of maternal body mass index and lipid levels in PE susceptibility. In contrast, fetal factors predisposing to PE were associated with increased risk of psychiatric disorders in adulthood, but positive effects on reproductive success. Collectively, these findings delineate distinct maternal and fetal genetic architectures underlying PE and highlight divergent biological pathways contributing to PE risk.

## Introduction

Hypertensive disorders of pregnancy (HDP) are a major cause of maternal and fetal morbidity and mortality^1^. Preeclampsia (PE), a life-threatening condition, is defined by maternal hypertension and proteinuria—or, in the absence of proteinuria, by other organ dysfunction—after 20 weeks of gestation. Both maternal and fetal factors are known to contribute to PE, globally affecting ∼4.6% [95% CI 2.7-8.2%] of pregnancies^2^. Known epidemiological risk factors of PE include higher maternal age and obesity, chronic hypertension, pregestational diabetes, kidney disease and autoimmune conditions^3^. Although the etiology of PE is complex, it is commonly associated with placental dysfunction^4^ and is known to have a substantial heritable component^5,6^.

Recent genome-wide association studies (GWAS) have provided insights into the biological basis of PE, showing that susceptibility involves a mix of cardiovascular regulators, including hypertension-related genes, as well as genes related to placental development and vascular remodeling^7–10^. The relationship of PE with hypertension and obesity-related traits such as body mass index (BMI) and lipid levels has been underscored by recent Mendelian Randomization studies^11,12^. A unique challenge in PE research, however, lies in the condition’s origin that is dependent on placenta and hence requires interaction of maternal and fetal tissues. Because mothers and children share roughly half of their genetic variants, disentangling maternal and fetal genetic contributions to PE remains challenging. This partial genetic overlap may also confound any causal inferences about the reasons for PE, and alleles influencing PE susceptibility are hypothesized to be under different selective pressures acting on maternal versus fetal interests^13–15^.

Here, we report a meta-analysis of GWAS on maternal and fetal genetic effects on PE, combining data from FinnGen R12^16^, FINNPEC^17^, Estonian Biobank (EstBB)^18^, and InterPregGen^7^, totaling 401,597 maternal and 435,076 fetal samples. We report 27 independent PE-associated lead variants (*P* < 5 × 10^−8^), including 12 novel and 3 fetal-specific loci. We provide detailed estimates of maternal and fetal genetic contributions to PE, demonstrating largely independent effects associated with distinct health outcomes. Leveraging causal inference via Latent Causal Variable (LCV) and Mendelian Randomization (MR) analyses, we show that only maternal genetic liability is closely linked to common risk factors for PE such as hypertension and BMI. In contrast, fetal genetic factors predisposing a mother to PE are associated with psychiatric risks in offspring. Collectively, our findings highlight independent and distinct maternal and fetal genetic architectures underlying PE.

## Results

### Identification of 23 maternal variants associating with preeclampsia

We included 401,597 maternal samples and 21,784 PE cases from FinnGen R12, FINNPEC, EstBB and InterPregGen to our meta-analysis, with ∼100,000 samples not included in our recent PE GWAS^8^ (Supplementary Table 1). We identified 23 independent loci associated with PE at genome-wide significance *P* < 5 × 10^−8^ (Table 1, Fig. 1). Eleven of these loci represent novel associations^8,9^, including signals near *PLEK (*rs34338164)*, LINC00243 (*rs915665)*, HLA-DRA (*rs3129882)*, VEGFA (*rs58414032)*, RP11-15B24.5 (*rs11139596)*, AKAP13 (*rs2241269)*, IL27 (*rs181207) and *PDE4C (*rs139078704). Three additional loci - *NPR3 (*rs1173727), *TNS2* (rs151036307) and *FURIN (*rs6224*) -* have been previously associated with HDP^8^. One maternal signal on chromosome 13 (*E1F4A1P7 / FLT1,* rs7318880) overlapped the known fetal PE locus^10^.

**Fig. 1.**
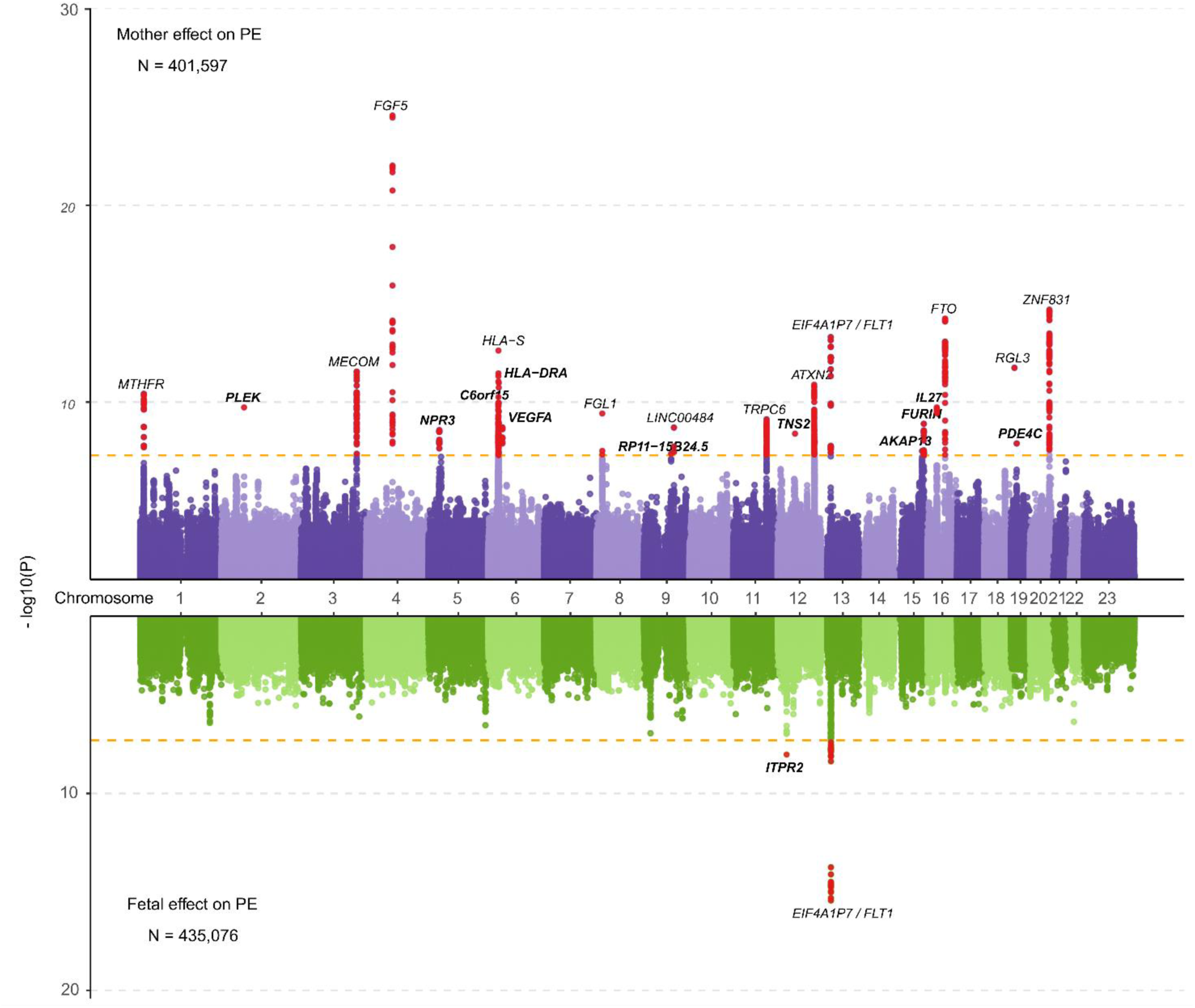
Summary of the preeclampsia (PE) GWAS meta-analysis results. A Miami plot showing maternal (purple shades) and fetal (green shades) associations with PE based on 21,784 maternal cases and 379,813 maternal controls, and 8406 fetal cases and 426,670 fetal controls. The plot indicates the −log_10_-transformed P value of each tested variant along the vertical axis and their chromosomal positions along the horizontal axis. The orange line signifies the genome-wide significance threshold of P = 5.0 × 10^−8.^ The genome-wide significant variants are indicated by red dots, with the nearest gene for each lead variant annotated. Genes residing in novel PE loci are shown in bold.

**Table 1.**
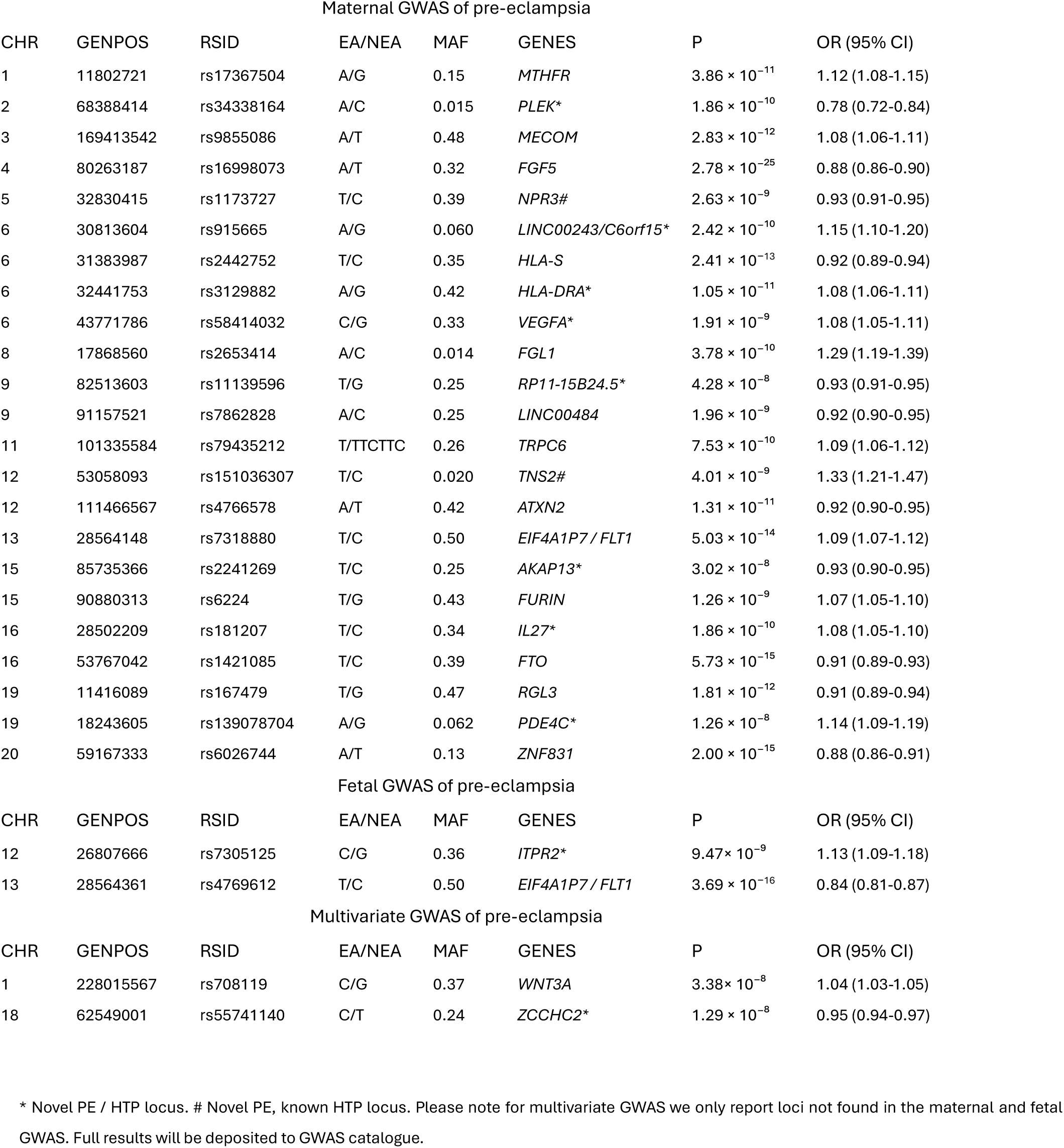
Lead variants in the genome wide association study (GWAS)-loci from Maternal, Fetal and Multivariate GWAS meta-analyses on preeclampsia (PE)

| Maternal GWAS of pre-eclampsia |  |  |  |  |  |  |  |
| --- | --- | --- | --- | --- | --- | --- | --- |
| CHR | GENPOS | RSID | EA/NEA | MAF | GENES | P | OR (95% CI) |
| 1 | 11802721 | rs17367504 | A/G | 0.15 | *MTHFR* | $3.86 \times 10^{-11}$ | 1.12 (1.08-1.15) |
| 2 | 68388414 | rs34338164 | A/C | 0.015 | *PLEK\** | $1.86 \times 10^{-10}$ | 0.78 (0.72-0.84) |
| 3 | 169413542 | rs9855086 | A/T | 0.48 | *MECOM* | $2.83 \times 10^{-12}$ | 1.08 (1.06-1.11) |
| 4 | 80263187 | rs16998073 | A/T | 0.32 | *FGF5* | $2.78 \times 10^{-25}$ | 0.88 (0.86-0.90) |
| 5 | 32830415 | rs1173727 | T/C | 0.39 | *NPR3#* | $2.63 \times 10^{-9}$ | 0.93 (0.91-0.95) |
| 6 | 30813604 | rs915665 | A/G | 0.060 | *LINC00243/C6orf15\** | $2.42 \times 10^{-10}$ | 1.15 (1.10-1.20) |
| 6 | 31383987 | rs2442752 | T/C | 0.35 | *HLA-S* | $2.41 \times 10^{-13}$ | 0.92 (0.89-0.94) |
| 6 | 32441753 | rs3129882 | A/G | 0.42 | *HLA-DRA\** | $1.05 \times 10^{-11}$ | 1.08 (1.06-1.11) |
| 6 | 43771786 | rs58414032 | C/G | 0.33 | *VEGFA\** | $1.91 \times 10^{-9}$ | 1.08 (1.05-1.11) |
| 8 | 17868560 | rs2653414 | A/C | 0.014 | *FGL1* | $3.78 \times 10^{-10}$ | 1.29 (1.19-1.39) |
| 9 | 82513603 | rs11139596 | T/G | 0.25 | *RP11-15B24.5\** | $4.28 \times 10^{-8}$ | 0.93 (0.91-0.95) |
| 9 | 91157521 | rs7862828 | A/C | 0.25 | *LINC00484* | $1.96 \times 10^{-9}$ | 0.92 (0.90-0.95) |
| 11 | 101335584 | rs79435212 | T/TTCCTC | 0.26 | *TRPC6* | $7.53 \times 10^{-10}$ | 1.09 (1.06-1.12) |
| 12 | 53058093 | rs151036307 | T/C | 0.020 | *TNS2#* | $4.01 \times 10^{-9}$ | 1.33 (1.21-1.47) |
| 12 | 111466567 | rs4766578 | A/T | 0.42 | *ATXN2* | $1.31 \times 10^{-11}$ | 0.92 (0.90-0.95) |
| 13 | 28564148 | rs7318880 | T/C | 0.50 | *EIF4A1P7 / FLT1* | $5.03 \times 10^{-14}$ | 1.09 (1.07-1.12) |
| 15 | 85735366 | rs2241269 | T/C | 0.25 | *AKAP13\** | $3.02 \times 10^{-8}$ | 0.93 (0.90-0.95) |
| 15 | 90880313 | rs6224 | T/G | 0.43 | *FURIN* | $1.26 \times 10^{-9}$ | 1.07 (1.05-1.10) |
| 16 | 28502209 | rs181207 | T/C | 0.34 | *IL27\** | $1.86 \times 10^{-10}$ | 1.08 (1.05-1.10) |
| 16 | 53767042 | rs1421085 | T/C | 0.39 | *FTO* | $5.73 \times 10^{-15}$ | 0.91 (0.89-0.93) |
| 19 | 11416089 | rs167479 | T/G | 0.47 | *RGL3* | $1.81 \times 10^{-12}$ | 0.91 (0.89-0.94) |
| 19 | 18243605 | rs139078704 | A/G | 0.062 | *PDE4C\** | $1.26 \times 10^{-8}$ | 1.14 (1.09-1.19) |
| 20 | 59167333 | rs6026744 | A/T | 0.13 | *ZNF831* | $2.00 \times 10^{-15}$ | 0.88 (0.86-0.91) |
| Fetal GWAS of pre-eclampsia |  |  |  |  |  |  |  |
| CHR | GENPOS | RSID | EA/NEA | MAF | GENES | P | OR (95% CI) |
| 12 | 26807666 | rs7305125 | C/G | 0.36 | *ITPR2\** | $9.47 \times 10^{-9}$ | 1.13 (1.09-1.18) |
| 13 | 28564361 | rs4769612 | T/C | 0.50 | *EIF4A1P7 / FLT1* | $3.69 \times 10^{-16}$ | 0.84 (0.81-0.87) |
| Multivariate GWAS of pre-eclampsia |  |  |  |  |  |  |  |
| CHR | GENPOS | RSID | EA/NEA | MAF | GENES | P | OR (95% CI) |
| 1 | 228015567 | rs708119 | C/G | 0.37 | *WNT3A* | $3.38 \times 10^{-8}$ | 1.04 (1.03-1.05) |
| 18 | 62549001 | rs55741140 | C/T | 0.24 | *ZCCHC2\** | $1.29 \times 10^{-8}$ | 0.95 (0.94-0.97) |

\* Novel PE / HTP locus. # Novel PE, known HTP locus. Please note for multivariate GWAS we only report loci not found in the maternal and fetal GWAS. Full results will be deposited to GWAS catalogue.

To investigate whether the identified maternal loci reflect shared genetic architecture with common risk factors for PE, we performed colocalization analysis using *coloc*^19^. Consistent with the known etiology of PE, 11 of the 23 maternal loci colocalized with causal variants for blood pressure, BMI, or autoimmune diseases (Supplementary Table 2). More broadly, 18/23 (78.2%) maternal loci showed evidence of association with hypertension (combined posterior probability (PP) > 0.8 for either the same or a different causal variant within the locus), 11/23 (47.8%) with BMI, and 11/23 (47.8%) with autoimmune diseases (Supplementary Table 2). Complementing these results, hypothesis-free functional annotation using FUMA^20^ showed that genes mapping to the maternal PE loci were significantly enriched in GWAS gene sets related to blood pressure, BMI, and inflammation-linked traits, as well as phenotypes associated with alcohol consumption and blood cell–related traits (Supplementary Table 3). Correspondingly, the maternal loci appeared significantly enriched to genes involved in hematopoiesis (*P*<1.8 × 10⁻^6^), and genes related to vasodilation (*P*=3.5 × 10⁻^5^) and blood pressure regulation (*P*=4.3 × 10⁻^5^), though the latter two failed to reach statistical significance after stringent Bonferroni correction (Supplementary Table 4). Finally, MAGMA tissue enrichment analysis identified uterus as the top tissue (*P*=0.010, Supplementary Fig. 1 and Supplementary Table 5), suggesting that PE-associated variants are preferentially located near genes with elevated uterine expression, though the enrichment was not statistically significant after Bonferroni correction. When querying the nearest genes in the loci for differential expression (DE) between normal and PE placentas^21^, we identified reduced *ATXN2* and *AKAP13* expression in PE placentas (adjusted *P*<0.05), in addition to previously reported increase in *FLT1*^9^ (Supplementary Table 6).

### Identification of fetal variants associated with maternal PE, including a signal at chromosome 12 near ITPR2

To evaluate the contribution of the fetal genome on maternal PE, we analyzed data from 435,076 samples, including 8406 cases. This meta-analysis extends the previous study by Steinthorsdottir et al. 2020^7^, by incorporating 52,929 additional Finnish samples (1631 cases) from FINNPEC^17^ and FinnGen R12^16^. We replicated the established fetal association at chromosome 13 (*EIF4A1P7* / *FLT1*), with lead variant rs4769612 (T/C, OR 0.84 [0.81-0.87], *P* = 3.7 × 10⁻^16^, showing the strongest association in the locus (Table 1, Fig. 1)^7,10^. A secondary independent PE variant within the same region (rs9508092 T/C, OR 1.16 [1.12-1.20], *P* = 7.5 × 10⁻^8^) narrowly missed the GWAS *P*-threshold. In addition, we report a novel fetal PE locus at chromosome 12 intronic to *ITPR2* (rs7305125, C/G, OR 1.13 [1.09-1.18], *P*=9.5 × 10⁻⁹). Beyond these two genome-wide significant signals, six additional loci showed suggestive associations (lead variant *P*<1.0 x 10^-6^ (Supplementary Table 7 and Supplementary Fig. 2).

A nearby *ITPR2* variant (rs11614652) was previously reported as nominally significant and followed-up by Steinthorsdottir et al.^7^ but did not reach genome-wide significance in that study. The potential candidate gene in the locus, *ITPR2,* encodes for inositol 1,4,5-trisphosphate receptor type 2, mediating intracellular calcium release. In humans, a loss-of-function in *ITPR2* has been associated with isolated anhidrosis, an inability to sweat despite normal sweat glands^22^, whereas in mice *Itpr2* function has been linked to promotion of cellular senescence^23^. Unlike the *EIF4A1P7 / FLT1* locus, which did not colocalize with hypertension variants, the *ITPR2* locus showed evidence for an independent hypertension-associate signal (PP>0.8, Supplementary Table 2) and overlaps GWAS associations for birth weight^24^ and cholesterol levels^25^. Given that only two loci reached genome-wide significance in the fetal analysis, FUMA annotation did not yield more insights into the data.

### Identification of two additional PE loci through multivariate GWAS

To identify additional variants influencing PE through either the maternal or fetal genome, or through both genomes, we performed a multivariate GWAS using genomic structural equation modeling (Genomic SEM)^26^. This approach jointly models summary statistics from the maternal and fetal GWAS to detect loci contributing to PE, irrespective of the genomic origin of the effects.

In the multivariate analysis we identified two additional variants that reached genome-wide significance, rs708119 at chromosome 1, intronic to *WNT3A* (C/G, OR 1.04 [1.03-1.05]*, P*=3.4× 10^−8^), previously associated with PE and gestational duration^9,27^, and rs55741140 on chromosome 18, intronic to *ZCCHC2* (C/T, OR 0.95 [0.94-0.97], *P*=1.3 × 10^−8^, Table 1, Supplementary Fig. 3). *WNT3A* is preferentially expressed in the placenta, plays a key role in placental development, and is an established DE gene between normal and PE placentas^9,21^, with prior studies suggesting its influence on PE risk may act through the fetal genome^9,28^. Resembling *WNT3A* function in cell proliferation and migration, *ZCCHC2* function has been linked to suppression of cell proliferation by inhibiting *MYC* activity, although the gene has not been implicated in PE in previously^23^. Colocalization analysis indicated that both SNPs rs708119 (*WNT3A*) rs57741140 (*ZCCHC2*) are unlikely to directly associate with hypertension, BMI or autoimmune diseases. However, closer inspection of the loci suggested that both loci contain overlapping GWAS associations with hypertension^29^, with additional associations to traits ranging from body size (*WNT3A*)^30^ to educational attainment (*ZCCHC2*)^31^.

### Power analysis

To evaluate the statistical power of the maternal and fetal GWASs to capture genetic effects on PE, we constructed a custom code to perform power simulations (Supplementary Code). Power was modeled under the assumption that mothers and children share 50% of their genome, across minor allele frequencies (MAF) from 0.01 to 0.5, odds ratios (OR) from 1.05 to 1.35, and case/total sample sizes of 21,784/401,597 for the maternal and 8406/435,076 for the fetal GWAS (Fig. 2a&b).

**Fig. 2.**
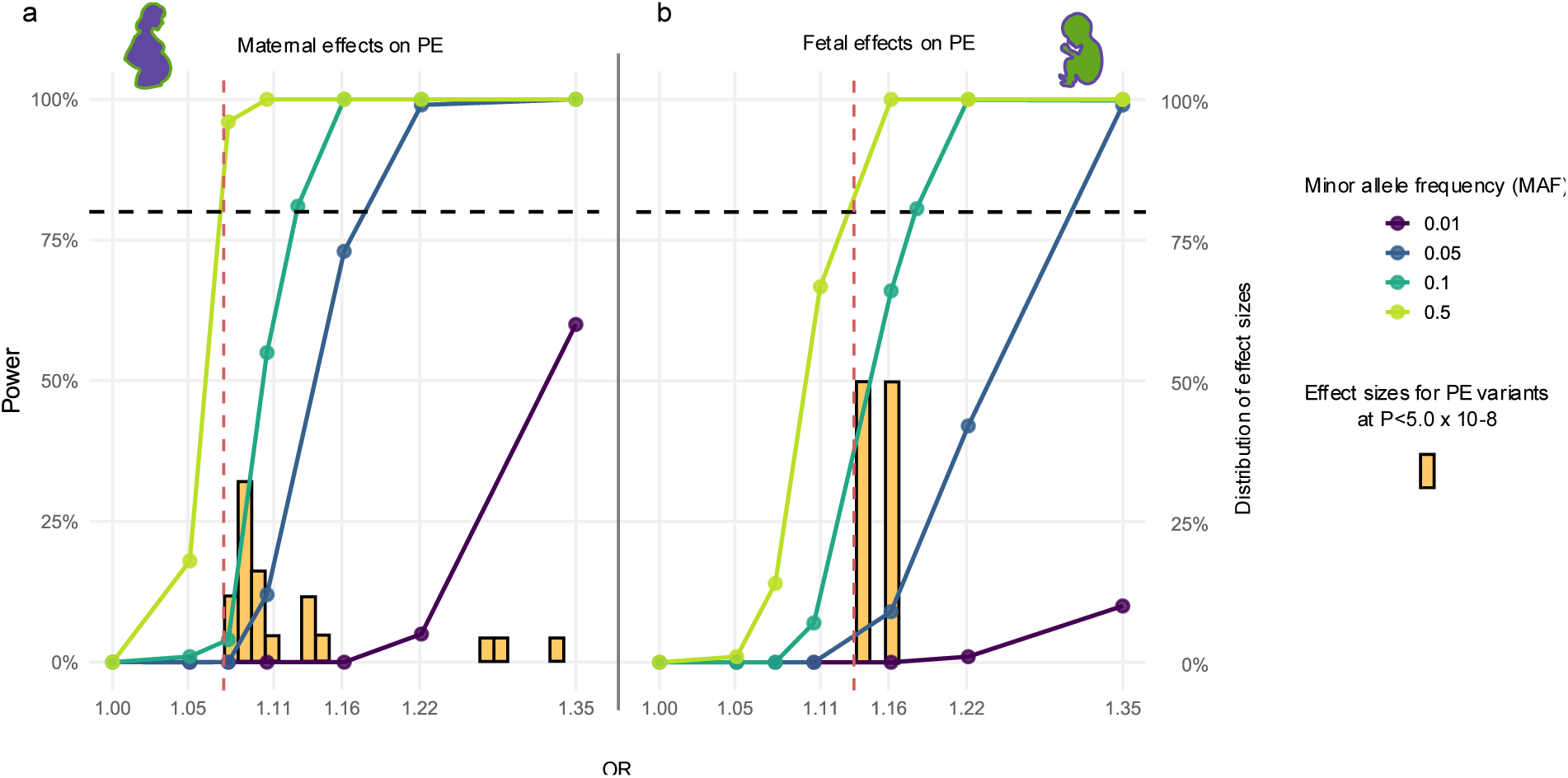
Power analysis. The plots illustrate maternal (a) and fetal (b) power curves to associate variants at genome-wide significance threshold (*P*<5e-08) with PE in this study. The maternal power curves are based on simulations of 401,597 samples and 21,784 PE cases, the fetal power curves on simulations of 435,076 samples and 8406 cases. Light green = MAF 0.5. Green = MAF 0.1. Blue = MAF 0.05. Dark purple = MAF 0.01. Yellow bars = distribution of observed effect sizes for lead SNPs. Black horizontal dashed line = 80% power threshold. Red vertical dashed line = smallest observed effect size for a lead SNP in the maternal/fetal analysis. MAF=minor allele frequency, PE=preeclampsia.

Assuming the variants identified in the maternal GWAS would hold only maternal effects, ≥80% power was achieved for OR >1.07 at MAF 0.5, OR >1.13 at MAF 0.1, and OR >1.18 at MAF 0.05. We had less power to detect fetal effects requiring OR >1.13 at MAF 0.5, OR >1.18 at MAF 0.1, and OR >1.30 at MAF 0.05 to reach the same threshold. Consistent with this power difference, 18/23 (∼78%) maternal variants displayed smaller effect sizes on PE than the two variants detected in the fetal GWAS. Notably, the smallest effect sizes detected in the GWASs were almost exactly those predicted by the power simulations to reach 80% power (Fig. 2a&b).

Based on the power analyses and predicted distributions of effect sizes in GWAS of complex disease^32^, we thus had reasonable power to detect common variants with relatively large effects in the maternal PE GWAS, but the current sample sizes may still limit the discovery of fetal PE variants, let alone variants with paternal effects (Supplementary Figure 4).

### Most SNP-based heritability of PE is maternal in origin, and maternal and fetal genetic effects are largely independent

Twin and family studies have shown that the overall heritability of PE is substantial, with maternal genetic factors explaining roughly 35% and fetal factors around 20% of variance in risk^6,7^. To quantify the contribution of common variants, we estimated SNP-based heritability (*h^2^* SNP) using genomic SEM, based on the maternal and fetal GWAS meta-analyses.

We observed a strong maternal genetic contribution to PE, with less evidence for fetal genetic effects, consistent with previous reports^6,7^. In total, maternal SNPs explained 11.9% of variance in PE risk, compared to 3.9% explained by fetal SNPs (Table 2). Given that mothers and children share 50% of their alleles, the high genetic correlation between maternal and fetal PE GWASs (*r_g_*=0.90 ±0.12, *P*=6.4e-14), needs considering when interpreting these estimates. After accounting for this genetic sharing between a mother and a fetus, 90.4% of the total SNP-based heritability appeared maternal in origin, still corresponding to 11.9% explained variance in PE, whereas fetal SNPs tagged only 1.3% of variance in maternal PE risk (Table 2).

**Table 2.** SNP-based heritability of preeclampsia (PE) and genetic correlations of maternal and fetal genome wide association studies (GWASs)

**Heritability (LDSC)**
| GWAS | Heritability (h<sup>2</sup> SNP) | P (h<sup>2</sup> SNP) |
| --- | --- | --- |
| Maternal effects on PE | 0.119 (0.010) | **6.1e-32** |
| Fetal effects on PE | 0.039 (0.012) | **0.0010** |

**Heritability, partitioned to maternal and fetal components (Genomic SEM)**
| Latent trait | Genetics from | Explained covariance | P |
| --- | --- | --- | --- |
| PE | Maternal+Fetal | 0.119 (0.015) | **1.8e-15** |
| Mother's PE | Maternal+Fetal | 0.013 (0.018) | 0.419 |
| PE~Mother's PE | Maternal+Fetal | -0.003 (0.013) | 0.786 |

**Genetic correlation (LDSC) and causal relationship (LCV)**
| Trait A | Trait B | $r_g$ | P ($r_g$) | GCP | GCP P |
| --- | --- | --- | --- | --- | --- |
| Maternal effects on PE | Fetal effects on PE | 0.90 (0.12) | **6.4e-14** | -0.61 (0.34) | **0.0079** |
| Mat PE, WLM corrected | FetPE, WLM corrected | 0.25 (0.31) | 0.42 | -0.68 (0.20) | **9.4e-06** |

The table shows liability-scale heritability estimates of PE based on GWAS results from LDSC and Genomic SEM models, with genetic covariance between maternal and fetal effects indicated. The genetic correlations and estimated causal relationships between the maternal and fetal effects, based on LDSC and LCV analyses, are shown. LDSC=Linkage Disequilibrium Score Regression. LCV = Latent Causal Variable model. WLM=weighted linear model. GCP=Genetic causality proportion.

Importantly, maternal and fetal genetic effects on PE were largely independent (genetic covariance = -0.003; *P* = 0.79), suggesting that different genetic and biological mechanisms in maternal and fetal tissues may contribute to PE pathophysiology. Using the Latent Causal Variable (LCV) method, we, however, plausibly observed evidence consistent with a causal influence of fetal genetic factors on maternal PE risk, but not the reverse (GCP -0.61 ±0.34, *P*=0.0079) (Table 2).

### Adjusting the PE GWAS results for fetal and maternal effects genome-wide

The baseline summary statistics from maternal and fetal GWAS are not adjusted for the 50% correlation between maternal and fetal effects at each locus. Therefore, the interpretation of maternal effects on PE can be complicated by fetal effects of the variants, and vice versa, as previously shown for the chromosome 13 *EIF4A1P7* / *FLT1* locus. To address this, we applied weighted linear model (WLM) framework, introduced by Warrington et al 2019^24^, to provide adjusted PE GWAS summary stats for maternal and fetal effects on PE. The WLM method linearly transforms GWAS summary stats effect estimates, accounting for allele sharing between mothers and fetus to disentangle the maternal and fetal genetic components.

After WLM adjustment, the genetic correlation between the maternal and fetal effects on PE (*r_g_*=0.25 ±0.31, *P*=0.42) was no longer significant and substantially lower than the unadjusted correlation, indicating effective separation of the two components (Table 2). To further validate the method, we compared WLM-adjusted estimates with directly estimated maternal and fetal effects for the 27 PE lead variants in 52,775 mother-child pairs from FinnGen and FINNPEC, derived from linear regressions including both maternal and fetal genotype for each variant. The adjusted estimates from these regressions showed strong concordance with WLM-correct estimates (*R*=0.77, *P*=1.3 × 10⁻^5^ for maternal, and *R*=0.99, *P*<2 × 10⁻^16^ for fetal SNP effects), confirming the accuracy of the correction (Figure 3 a&b, and Supplementary Table 8).

**Fig. 3.**
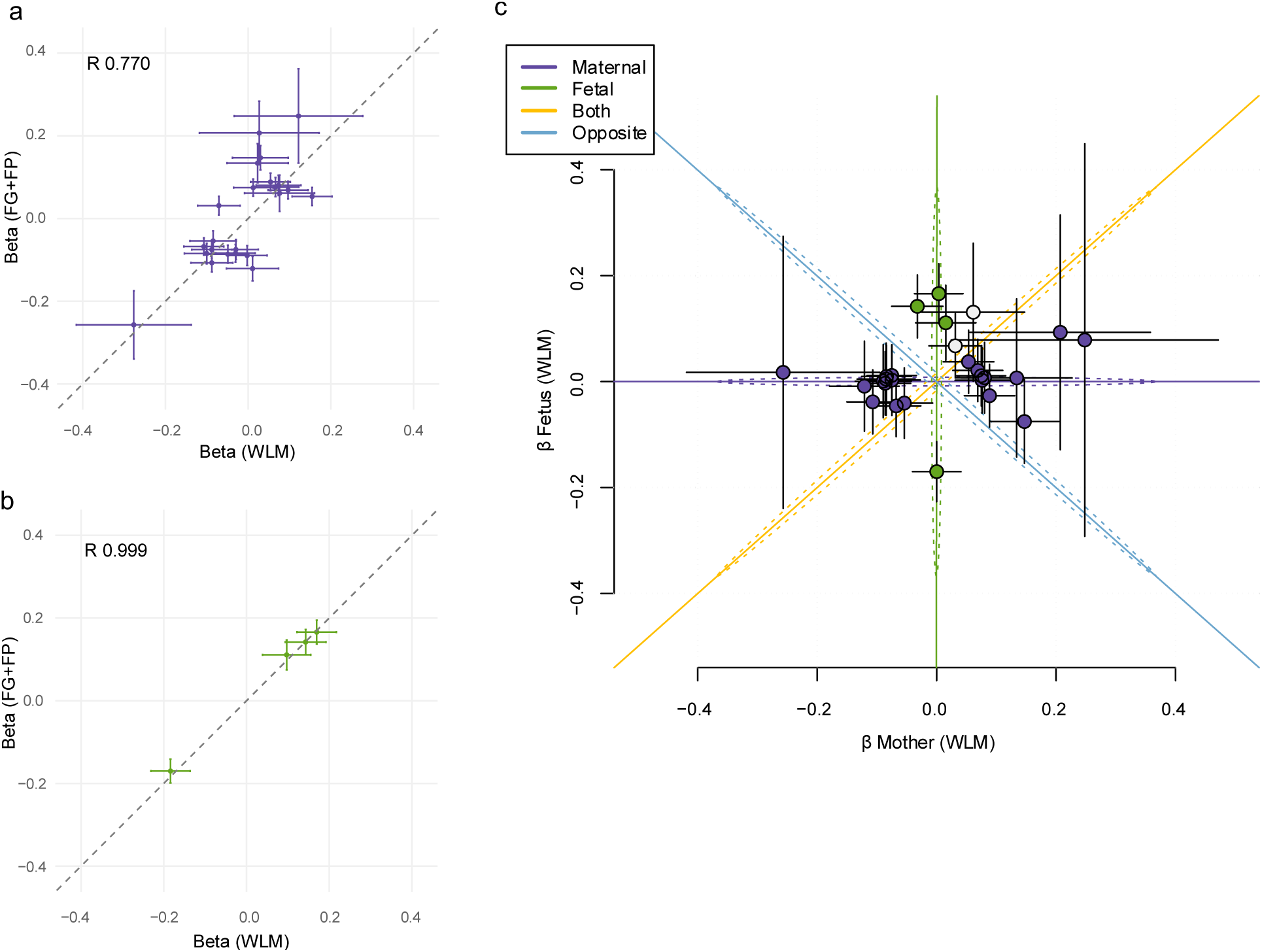
Separating maternal and fetal effects of preeclampsia (PE) lead variants. Panels a and b show correlation between the PE lead variants’ WLM-corrected and observed effects on PE, for mothers (a, purple) and children (b, green). For panels a and b the x-axis show the WLM corrected beta of original summary stats, and the y-axis observed effects in FinnGen (N=51,215, N cases = 829) and FINNPEC (N=1560, N cases=739) mother-child pairs, from a linear model taking into account both maternal and fetal genotype. Panel c illustrates division of the identified lead variants into maternal and fetal categories using *linemodels,* based on the variants’ WLM corrected effect sizes. Variants with posterior probability (PP>0.9) for maternal effects are coloured purple, and variants with PP>0.9 for fetal effects are colored green. The variants with PP<0.9 for either maternal or fetal effects are left grey.

Although the WLM is expected to give more accurate maternal and fetal effect estimates for the studied variants, the downside of the correction is that meanwhile it also increases the uncertainty of the estimates in the form of increased standard errors. Consequently, the proportion of variance in PE explained by the WLM-corrected maternal data was reduced to 3.0% but stayed significant (*P*=1.0 x 10^-7^), while the adjusted fetal data no longer showed significant heritability (*P*=0.262, Table 2).

### Allocation of identified variants into maternal and fetal loci

To formally assign the identified loci to maternal and fetal categories, we applied *linemodels*^33^ to the WLM-corrected GWAS results (Table 3 and Fig. 3C). *linemodels* allows for probabilistic clustering of variables based on their observed effect sizes on two outcomes, in this case maternal and fetal effects on PE.

**Table 3.** Annotation of variants to maternal and fetal loci.

| CHR | SNP | Gene | Orig. GWAS | PP-Mat | PP-Fet | PP-Both | PP-Opp. | Adj. $\beta$ mat. | Adj. $\beta$ fet. |
| --- | --- | --- | --- | --- | --- | --- | --- | --- | --- |
| **Maternal variants** |  |  |  |  |  |  |  |  |  |
| 1 | rs17367504 | MTHFR | Maternal | **0.961** | 0 | 0 | 0.039 | 0.147 | -0.075 |
| 2 | rs34338164 | PLEK | Maternal | **0.979** | 0.004 | 0.003 | 0.014 | -0.257 | 0.046 |
| 3 | rs9855086 | MECOM | Maternal | **0.992** | 0 | 0.004 | 0.004 | 0.075 | 0.011 |
| 4 | rs16998073 | FGF5 | Maternal | **1** | 0 | 0 | 0 | -0.107 | -0.037 |
| 5 | rs1173727 | NPR3 | Maternal | **0.990** | 0.001 | 0 | 0.009 | -0.075 | 0.012 |
| 6 | rs915665 | LINC00243 | Maternal | **0.982** | 0.010 | 0.002 | 0.006 | 0.134 | 0.024 |
| 6 | rs3129882 | HLA-DRA | Maternal | **0.987** | 0.003 | 0.008 | 0.002 | 0.069 | 0.014 |
| 6 | rs58414032 | VEGFA | Maternal | **0.990** | 0.005 | 0.002 | 0.003 | 0.076 | 0.000 |
| 6 | rs2442752 | HLA-S | Maternal | **0.997** | 0 | 0.002 | 0.001 | -0.086 | -0.003 |
| 8 | rs2653414 | FGL1 | Maternal | **0.955** | 0.019 | 0.020 | 0.006 | 0.207 | 0.111 |
| 9 | rs11139596 | RP11-15B24.5 | Maternal | **0.992** | 0.002 | 0.003 | 0.003 | -0.075 | 0.003 |
| 9 | rs7862828 | LINC00484 | Maternal | **0.992** | 0.001 | 0 | 0.007 | -0.085 | 0.005 |
| 11 | rs79435212 | TRPC6 | Maternal | **0.991** | 0.002 | 0.003 | 0.004 | 0.080 | 0.007 |
| 12 | rs4766578 | ATXN2 | Maternal | **0.998** | 0 | 0 | 0.002 | -0.084 | 0.010 |
| 12 | rs151036307 | TNS2 | Maternal | **0.907** | 0.063 | 0.02 | 0.010 | 0.248 | 0.079 |
| 15 | rs2241269 | AKAP13 | Maternal | **0.901** | 0.058 | 0.037 | 0.004 | -0.054 | -0.040 |
| 15 | rs6224 | FURIN | Maternal | **0.920** | 0.030 | 0.049 | 0.001 | 0.053 | 0.037 |
| 16 | rs181207 | IL27 | Maternal | **0.994** | 0 | 0 | 0.006 | 0.089 | -0.019 |
| 16 | rs1421085 | FTO | Maternal | **0.934** | 0.007 | 0.057 | 0.002 | -0.068 | -0.048 |
| 19 | rs167479 | RGL3 | Maternal | **0.995** | 0 | 0.001 | 0.004 | -0.089 | 0.001 |
| 20 | rs6026744 | ZNF831 | Maternal | **0.998** | 0 | 0.002 | 0 | -0.121 | -0.007 |
| **Fetal variants** |  |  |  |  |  |  |  |  |  |
| 12 | rs7305125 | ITPR2 | Fetal | 0 | **0.993** | 0 | 0.007 | -0.032 | 0.142 |
| 13 | rs7318880 | EIF4A1P7 | Maternal | 0 | **1** | 0 | 0 | 0.006 | 0.161 |
| 13 | rs4769612 | EIF4A1P7 | Fetal | 0 | **1** | 0 | 0 | -0.002 | -0.165 |
| 18 | rs55741140 | ZCCHC2 | Multivariate | 0.046 | **0.949** | 0.003 | 0.002 | 0.015 | 0.112 |
| **Unassigned variants** |  |  |  |  |  |  |  |  |  |
| 1 | rs708119 | WNT3A | Multivariate | 0.427 | **0.509** | 0.052 | 0.012 | 0.031 | 0.068 |
| 19 | rs139078704 | PDE4C | Maternal | **0.502** | 0.438 | 0.057 | 0.003 | 0.061 | 0.131 |

The table shows the posterior probability (PP) for variants acting through the maternal or fetal genome. The annotations are based on WLM-adjusted data, with betas shown. The model with the highest PP for each SNP is indicated in bold. Allocation to maternal/fetal/unassigned variants was based on PP>0.9.

Among the 23 genome-side significant maternal variants, for 21 (91%) we detected strong statistical support (PP>0.90) for these variants acting exclusively through the maternal genome (Table 3 and Fig. 3C). In contrast, both variants detected in the fetal GWAS, rs4769612 (*EIF4A1P7 / FLT1*) and rs7305125 (*ITPR2*) were classified to act only through the fetal genome. Notably, *linemodels* also identified rs55741140 (*ZCCHC2)*, discovered in the multivariate GWAS, as acting through the fetal genome.

For two variants - rs139078704 (*PDE4C*) and rs708119 (*WNT3A*) - the model was inconclusive. However, in both cases, the adjusted fetal effect sizes were approximately twice as large as the maternal estimates, consistent with predominant fetal contributions. The potential fetal association at *WNT3A* aligns with an earlier report suggesting fetal effects at this locus^9^. Collectively, these analyses add support to the concept that most of the PE risk is mediated by the maternal genome, whereas a smaller subset of variants still represent independent fetal effects, underscoring that maternal and fetal effects on PE are largely uncoupled at the genetic level.

### Genetic correlations

To gain insight into the broader genetic architecture of PE, we next estimated genetic correlations of both the maternal and fetal GWASs with 40 selected complex traits representing a broad range of phenotype categories linked to PE etiology, including anthropometric, biomarker, cardiometabolic and neurological endpoints. Given the substantially lower heritability estimates for the WLM-corrected data, that may limit both genetic correlation and causal inference, we focused on reporting the findings based on the unadjusted maternal and fetal GWAS results. We, however, provide similar estimates also for the WLM-corrected datasets.

For the maternal PE GWAS, LDSC analysis revealed multiple expected and previously reported correlations, including positive genetic overlap with hypertension, cardiovascular diseases, and type 2 diabetes (T2D)^7^ (Fig. 4A and Supplementary Table 9). In addition, we noted statistically significant (*P*<0.00125) positive genetic correlations e.g. to higher body fat content (*r_g_*=0.27±0.07, *P*=0.00017), hypothyroidism (*r_g_* 0.28±0.07, *P*=6.4 x 10^-5^), non-alcoholic fatty liver disease (NAFLD) (*r_g_* 0.35±0.10, *P*=0.00032) and poorer overall health (*r_g_* -0.20±0.05), *P*=7.9 x 10^-5^, Fig. 4A and Supplementary Table 9). The fetal GWAS on PE, however, did not show statistically significant correlations to the studied traits, except in case of hypertension (*r_g_* 0.46±0.13, *P*=2.3 x 10^-5^, Fig. 4A and Supplementary Table 9). This result likely stemmed from the unadjusted fetal GWAS carrying over some maternal effects of the SNPs, as shown by the correlation disappearing in the WLM-adjusted fetal data (Supplementary Table 9). Overall, the genetic correlation patterns, however, were largely consistent between the original and WLM-adjusted datasets (Supplementary Fig.4 & Supplementary Table 9).

**Fig. 4.**
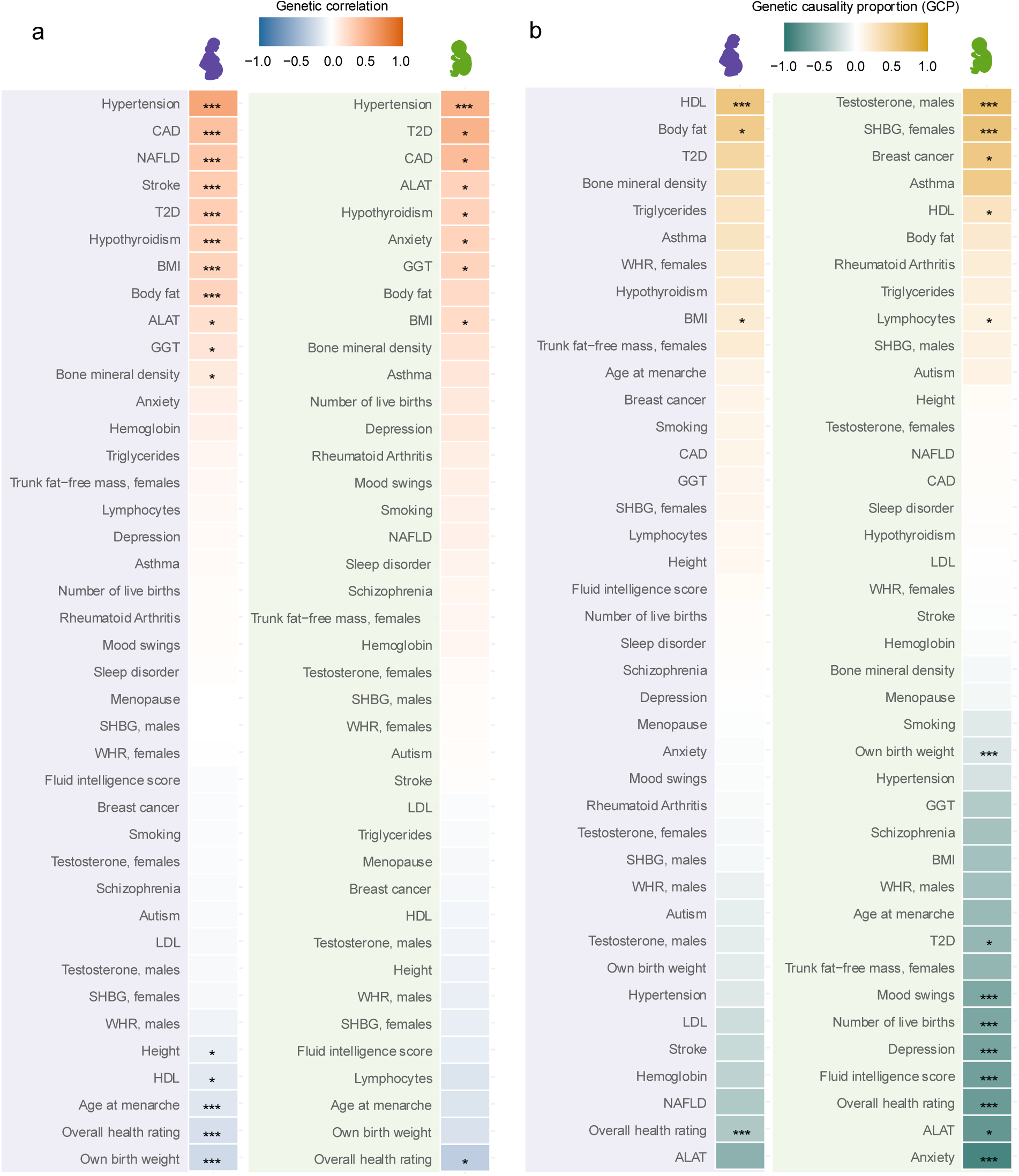
Genetic correlations and causality between maternal and fetal effects on preeclampsia (PE) and selected traits. The panel (a) shows genetic correlations, and panel (b) inferred causality from latent causal variable (LCV) analyses. Purple shadowing = results for maternal genome wide association study (GWAS). Green shadowing = results for fetal GWAS. Blue=negative genetic correlation, red=positive genetic correlation. Yellow = Causality of named traits on PE, Dark green = Causality of PE on the named traits. * *P*<0.05, *** *P*<0.00125, corresponding for conservative Bonferroni correction of 40 independent traits.

### Causality

While genetic correlations quantify the extent of shared genetic architecture between traits, they do not imply causation. To explore potential causal relationships between the 40 selected traits and the maternal and fetal genetic effects on PE, we applied Latent Causal Variable (LCV) analysis^34^ (Fig. 4B and Supplementary Table 9). The advantages of the LCV framework are that it exploits genome-wide summary statistics to infer causal relationships, making it particularly suitable for traits with relatively few genome-wide significant loci, like in the case of fetal effects on PE. In addition, LCV gives a clear indication of both the strength and the direction of causality (whether trait 1 is causal to trait 2 (indicated by positive GCP-value) or vice versa (indicated by negative GCP-value)), whilst being also much more resistant to type I errors caused by genetic correlation and pleiotropy compared to conventional MR. For comparison, we provide also conventional MR estimates from TwoSampleMR^35^ (Supplementary Table 10 and Supplementary Fig. 6&7).

Consistent with the genetic correlation results, the causality analyses highlighted that maternal and fetal pathways contributing to PE are largely distinct (Fig. 4A&B). We recapitulate the findings that higher BMI^11^ (genetic causality proportion (GCP) 0.21, *P*=6.3x10^-3^), and lower HDL levels^12^ in mothers (GCP 0.60, *P*=9.4x10^-11^) can work as partial causal factors to PE. In addition, genetic predisposition to higher body fat (GCP 0.53, *P*=0.047) was nominally associated with PE, whereas a PE diagnosis appeared causal for lower overall health rankings in females (GCP -0.38, *P*=9.3x10^-6^, Fig. 4B and Supplementary Table 9). In contrast, there was less support for genetic predisposition to these traits being a causal factor for being born to a PE-pregnancy, though we noted that lower genetically determined male testosterone (GCP 0.52, *P*=9.4x10^-11^) and female SHBG levels in fetuses (GCP 0.68, *P*=1.9x10^-7^) could promote fetal PE, suggesting intrauterine hormone levels as potential PE risk factors (Fig. 4B and Supplementary Table 9).

Finally, the LCV analyses indicated that fetal genetic risk for PE links not only to lower birth weight (GCP -0.33, *P*=3.4x10^-11^), but also associates with several psychiatric traits, including anxiety, depression and mood instability (GCP -0.90, -0.70 and -0.69, respectively, all *P*<1.0x10^-9^). Fascinatingly, despite these risks and suggesting potential reasons for maintaining fetal PE susceptibility alleles in population, fetal PE risk was positively associated with number of live births (*r_g_*=0.141, GCP -0.63 (0.19), *P*=4.2 x 10^-12^). Intrigued by this finding, we tested for fetal PE effects also on number of children fathered (NCF) and found a similar subtle but positive effect (*r_g_*=0.091, GCP -0.76 (0.18), *P*=4.1 x 10^-22^) though only in the WLM-corrected data. Collectively, these findings indicate that whereas maternal metabolism related factors can act as direct causal drivers of PE, fetal genetic susceptibility to PE can increase the child’s risk of adverse psychiatric outcomes later in life, balanced by potential positive effects on the child’s future reproductive success.

## Discussion

In this study, we sought to identify and disentangle maternal and fetal genetic contributions to PE. Altogether we identified 27 variants, of which four were fetal, showing genome-wide significant associations with the mother’s risk of PE. Although most statistically significant associations were maternal in origin, our analyses were consistent with prior evidence indicating that both maternal and child genomes contribute to PE susceptibility. The divergence between maternal and fetal effect estimates underscores the value of explicitly modeling both genomes to avoid confounding and to reveal distinct etiological pathways for PE. While our results support the concept that PE might be a first sign of a mother’s lifetime cardiovascular disease risk and overall poorer health^8,36,37^, we also provide evidence that being born from a PE pregnancy may contribute to several adverse health outcomes in the child, particularly psychiatric comorbidities.

In the maternal data, the strongest associations were observed at loci previously linked to blood pressure regulation and vascular function, reinforcing the hypothesis that maternal cardiovascular physiology is a central determinant of PE risk, and that PE can be the first sign of an increased risk for cardiac events in the future^8,36,38,39^. However, instead of pointing impaired blood pressure regulation as the direct causal culprit for PE^40^, our genetic evidence rather points toward obesity - potentially leading to elevated blood pressure and poorer metabolic health - in determining a mother’s PE risk at a population level. In turn, a PE event appeared causal for later poorer overall health, as self-reported by 40-70 yrs old women from the UK Biobank. These findings complement previous MR studies, suggesting higher BMI and lower HDL levels as causal factors to PE^11,12^. We noted that some of the genetic variants that predispose to PE correlate with adverse cardiometabolic events such as CAD and stroke later in life, supporting recent findings^41^. Our study, however, did not imply PE as a direct causal factor for heart disease, instead suggesting that the relationships between PE and cardiometabolic health results from genetic pleiotropy – i.e. shared genetic factors independently contributing to PE and cardiovascular disease risks. Finally, the discovery of several immune-system related genetic loci in the maternal GWAS, many residing in the major histocompatibility (MHC) complex located on chromosome 6, pointed the maternal immune system as a potential contributor to PE, aligning with biological models where impaired placentation and maladaptation of maternal–fetal immune interactions contribute to disease pathogenesis^4,38,42^. Yet we observed no direct causality between studied autoimmune disease and PE, supporting a model where general immune system function rather than autoimmune pathology per se is more closely linked with PE progression.

Epidemiological studies have shown that people born from a PE pregnancy are at increased risk for psychiatric diagnoses ^38,43–45^. Here we demonstrate that only the fetal, but not maternal, genetic profile predisposing to PE associates with increased risk of psychiatric disease. One potential interpretation is that fetal genes increasing PE susceptibility could promote psychiatric disease risk later in life. Alternatively, a PE event with altered intrauterine conditions and associated postnatal complications, could directly affect developmental trajectories of the child. Intriguingly, prior studies have shown that presence of postnatal complications following PE pregnancies is most strongly associated with a child’s psychiatric disease risk^43^. Addressing the exact mechanism linking fetal predisposition to PE and psychiatric diseases remains an important avenue for research in the future. Nonetheless, while current evidence supports a causal contribution of PE to elevated psychiatric disease risk among exposed children, the absolute probability of any given individual receiving such a diagnosis remains modest.

Our heritability analyses indicated that most of the common genetic risk for PE can be attributed to maternal factors, which is in line with earlier studies^7^. The current data explain roughly 12% of the maternal genetic heritability of PE, enabling well-powered polygenic score (PGS) analyses and supporting robust estimation of genetic correlations and causal relationships with other traits. However, after considering the maternal part, common genetic variation explained much less about the fetal contribution to PE (∼1%). Since substantial fetal contribution to PE has been estimated in genetic studies^5–7,10^, this likely reflects limited power in the fetal GWAS. According to some estimates, for many complex diseases, hundreds of thousands of cases would be required to explain 50% of the heritability^46^. Indeed, our power analyses indicate that detecting fetal genetic effects on PE remains underpowered under current GWAS sample sizes, posing a problem that could be only resolved by global collaborative efforts^32,46^. Extending these speculations, detecting any paternal genetic effects on PE in GWAS settings - while considering potential mediation through fetal genetics - would require genotyped and phenotyped trio datasets of similar size as used in this study, which are not currently available.

The results of this study raise ground for speculation about the reasons for the persistence of PE susceptibility alleles in human populations. From an evolutionary perspective, variants that increase PE risk through maternal or fetal pathways may persist due to balancing selection between maternal and fetal interests^47^. Notably, many maternal PE variants are related to maternal obesity, whereas obesogenic environment has not persisted that long^48,49^. While obesity-linked alleles may predispose mothers to hypertension and PE, they may enhance fetal nutrient acquisition or indicate metabolic economy conferring fitness benefits in pre-modern environments. Our data interestingly suggest that fetal alleles increasing PE susceptibility might promote later reproductive success of the child, despite PE associated risks for mother and child during childbirth, and e.g. potential adverse psychiatric outcomes for the child. Overall, these observations are in line with theories of evolutionary trade-offs between the fetus and the mother during pregnancy, where the energetic demands of the fetal brain and deeply invasive placenta, features unique to humans, contribute to PE’s prevalence^14,15,47^. The finding of genetically distinct and largely uncorrelated maternal and fetal genetic effects on PE in this study supports the idea that opposing evolutionary pressures may have shaped PE genetics, consistent with the principle that genetic factors exerting opposite effects on fitness in different groups tend to show low genetic correlation^50^.

In conclusion, our results demonstrate that the maternal and fetal effects on PE are largely uncoupled at the genetic level, and associate with different health risks. Whereas effective preventive measures of PE should primarily target the expectant and prospective mothers, children born from PE pregnancies would instead benefit from extended support and monitoring during the postnatal period. Overall, our study thus provides new insights into the distinct genetic mechanisms underlying maternal and fetal contributions to PE, highlighting opportunities for improved prevention and targeted intervention.

## Methods

### Cohorts and phenotypes

The GWAS meta-analysis on maternal and fetal effects on PE totaled 401,597 maternal and 435,076 fetal samples, from FinnGen R12, FINNPEC, EstBB and InterPregGen. For the meta-analysis, we combined GWAS data from FinnGen Release 12 (R12)^16^, FINNPEC^17^ and EstBB^8,18^ data, together with publicly available InterPregGen GWAS meta-analysis results^7^. The final meta-analyses data included ∼100,000 more samples than the most recent maternal PE GWAS from our group^8^ (Supplementary Table 1), and ∼52,000 more fetal samples than recent meta-analysis by Steinthorsdottir et al.^7^ For InterPregGen and EstBB details of the participating cohorts and patient inclusion criteria have been described earlier^7,8^. In short, for all participating cohorts and datasets (FINNPEC, FinnGen, EstBB and InterPregGen) PE diagnoses were either independently verified according to international criteria (e.g. FINNPEC) or obtained based on ICD codes recorded by a clinician (e.g. FinnGen, EstBB). FinnGen ICD codes come from Finnish registers which show substantial concordance with diagnoses independently verified from medical records^44^. All required ethical approvals were obtained from international, national or regional ethics committees as described in detail in Supplementary Information.

### Association analyses

The GWAS analyses for maternal and fetal effects on PE in FinnGen and FINNPEC were performed with regenie v2.2^51^ under a logistic model. Analysed SNPs were restricted to variants with MAF ≥ 0.1 % and imputation quality ≥0.6. We included both autosomal and X-chromosomal variants into the analysis. X-chromosomal effects in male fetuses were based on (0,2) allele dosage coding. First 10 Principal Components, mother birth year, and genotyping batch (FinnGen) were used as quantitative covariates in the maternal runs. For fetal analyses we in addition included fetal sex, mother age, and own birth year as covariates. For FinnGen analyses, we included only parous women, and excluded all women sampled from the FINRISK-cohorts, since these were included in the InterPregGen meta-analysis^7^, resulting in 192,273 women with 9296 PE diagnoses (O15_PRE_OR_ECLAMPSIA) to be included. The details of the association runs in EstBB and InterPregGen have been described before^7,8^. The EstBB and InterPregGen summary statistics were lifted to human genome build 38 before the meta-analysis. An inverse variance–weighted meta-analysis, corrected for genomic inflation with METAL software^52^, was performed to produce the final summary statistics on maternal and fetal effects on PE. The meta-analyses included 21,360,942 maternal and 14,883,669 fetal variants. Lead SNPs were defined as independent variants with the smallest P-value within a 1MB window (±500kb from the lead variant). R2 threshold of 0.1 was used to define independent lead SNPs with P < 5 × 10^−8^ within the identified loci.

### Pathway, tissue enrichment and co-localization analyses

For formal co-localization analyses to assess whether PE and hypertension (I9_HYPTENS) ^16^, BMI^53^ and autoimmune disease (AUTOIMMUNE)^16^ share the same causal variant we used *coloc*^19^ in R with posterior probability (PP)>0.80 as colocalization criterion. Tissue and gene set enrichment analyses were carried out with MAGMA implemented in FUMA using default settings^20^. The GWAS catalog look-up was performed on version e110_r2023-07-20. To evaluate in which tissues the genes near GWAS loci were preferably expressed, we used the full distribution of SNP *P*-values and the GTEx v8 30 general tissue types. In the pathway analysis by MAGMA, the full distribution of SNP *P*-values was used to query MsigDB v2023.1Hs with 17023 genes sets, and a conservative Bonferroni correction was applied to the results. For differential expression of the identified genes in normal vs. PE placentas, we queried the online database of the human placenta transcriptome^21^.

### Multivariate GWAS

We performed a multivariate GWAS using GenomicSEM^26^ to jointly model maternal and fetal genetic effects on preeclampsia (PE). Maternal and fetal GWAS summary statistics were pre-processed and combined into an LD score regression–based genetic covariance matrix using the European 1000 Genomes reference panel. We then specified a structural equation model in which SNP effects were decomposed into maternal and fetal components, allowing both shared and distinct contributions to PE risk to be estimated. The model was fit genome-wide with the userGWAS function, producing SNP effect estimates and *P*-values for maternal and fetal contributions simultaneously, increasing power to detect genetic loci affecting PE risk while accounting for the genetic correlation between the fetal and maternal datasets.

### Power analysis

We designed simulations to estimate statistical power to capture maternal / fetal effects on mother’s PE with the available datasets. The R script designed to run the power analysis, partly modified based on power analysis from Leinonen et al 2024^54^ is available as Supplementary Code. For each Monte Carlo replicate (N=100 iterations per MAF/β combination), we first simulated genotypes for N = 401,597 / 435,076 samples under Hardy–Weinberg equilibrium (HWE), given a specified MAF (range 0.01-0.50). Fetal genotypes were generated directly from HWE expectations. Maternal genotypes were then simulated conditional on the fetal genotype to reflect ∼50% genetic correlation between mother and child. Specifically, one maternal allele was set equal to the allele transmitted to the child, while the second allele was sampled independently according to the MAF.

Phenotypes were generated under a logistic model in which case–control status was determined by a baseline prevalence (Ncases/N) and predefined effect sizes (β range 0.05-0.3) for maternal (βm) and offspring (βo) genotypes. For each replicate, case status was sampled from a Bernoulli distribution with probability equal to the modeled risk. We then fit a logistic regression model of PE status on maternal and fetal genotypes. We finally transformed the simulated βs to ORs for plotting to assist in interpretation of the results. Power was defined by the number of simulations where the maternal/fetal effect reached genome-wide significance (*P* < 5 × 10^−8^). For the trio analyses, we used the same principles to simulate paternal, maternal and fetal genotypes and maternal phenotypes (Supplementary Code).

### Heritability estimations

We applied LD score regression (LDSC)^55^ and Genomic Structural Equation Modeling (GenomicSEM)^26^ to estimate SNP-based heritability of preeclampsia (PE) and to partition this heritability into maternal and fetal components. Liability -scale heritability estimations, based on effective population sizes of 61,271 maternal and 21,447 fetal cases, were obtained from LDSC analyses. The proportion of preeclampsia (PE) SNP heritability, partitioned into maternal and fetal explained variances was estimated based on the LDSC results as follows:

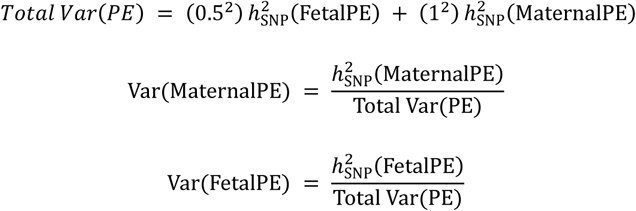

LDSC was used to generate genetic covariance matrices for maternal and fetal GWASs on PE. To formally distinguish the maternal and fetal genetic contributions to PE, we specified a user-defined structural equation model (SEM) in which PE risk was modeled as a function of maternal genetic effects, fetal genetic effects, and their covariance. Adjusted maternal and fetal SNP-heritability and the covariance between the maternal and fetal GWAS were then estimated by fitting this model to the LDSC-derived covariance matrix using the usermodel() function in GenomicSEM.

### Genetic correlations

Genetic correlations between maternal and fetal effects on PE and 40 phenotypes from broad trait categories were estimated using LDSC^55^, including autosomal variants only. The traits were selected on the basis of these being representative of the vast phenotype and disease classes that have been thought to either to contribute to or be caused by PE (referenced in Supplementary Table 8). The summary statistics for the 40 traits were downloaded directly from the source repositories and analyzed locally. The original sources are referenced in Supplementary Table 7. For genetic correlations, pre-computed LD Scores from 1000 Genomes Europeans, excluding the HLA region were used.

### Separating maternal and fetal effects

To obtain adjusted GWAS summary statistics for maternal and fetal effects on PE, we applied a Weighted Linear Model (WLM) introduced by Warrington et al. 2019^24^. WLM performs a linear transformation to the maternal and fetal GWAS summary statistics, weighted by the sampling variance, to generate corrected estimates that separately reflect direct fetal and direct maternal effects. The approach enables partitioning of genetic associations into maternal and fetal components using only GWAS summary data, without requiring individual-level genotype information. To assess the robustness of WLM correction in case of PE data, we compared the WLM adjusted estimates of the lead SNPs with direct effect estimates from genotyped FinnGen mother-child pairs with information on mother’s PE.

To probabilistically classify the identified lead variants to maternal and fetal categories, based on the WLM-corrected effect estimates, we applied the *linemodels* R package^33^ The method clusters variants into predefined linear relationship models between four effect dimensions (maternal, fetal, both, opposite). The expected magnitude of effects (scale) was set to 0.15 (corresponding to 95% effects being under logOR of 0.3), slopes were set to reflect the four different scenarios “maternal-only” (0), “fetal-only” (1000), “both effects” (1) and “opposite effects” (-1), with correlation values of 0.999. The *linemodels* algorithm uses observed effect estimates—and their standard errors and covariance—to compute posterior membership probabilities (PP) for each variant across these models. Variants were allocated to maternal, fetal, both and opposite categories if the PP exceeded 0.90^33^.

### Causality analyses

To investigate the potential causal relationships between maternal and fetal genetic liability to preeclampsia (PE) and the 40 traits subjected to genetic correlation analyses, we performed Latent Causal Variable (LCV) analysis^34^. We chose LCV as our primary causality method since it exploits genome-wide summary stats, that is particularly useful when the number of identified loci in GWAS is low like in case of the fetal effects on PE. Like LDSC and GenomicSEM, LCV uses genome-wide covariance structures and therefore can be used in contexts where some or many of the phenotypes are measured in the same people. In addition, LCV gives a clear indication of the direction of causality whilst being more robust to genetic pleiotropy compared to standard MR^34^. LCV estimates the proportion of genetic correlation between two traits that can be attributed to a causal relationship, summarized by the genetic causality proportion (GCP). A GCP near zero suggests that any genetic correlation would be mostly due to horizontal pleiotropy (a same genetic variant has independent effects on two traits), whereas a GCP approaching ±1 indicates strong evidence that genetic liability to one trait is partially causal for the other. LCV analysis was conducted in R using the genome-wide summary statistics as subjected to the LDSC analyses, restricted to well-imputed HapMap3 SNPs excluding the HLA-region, and LD scores computed from the European 1000 Genomes reference panel.

As a complementary approach, we conducted standard two-sample Mendelian randomization (MR) using the TwoSampleMR R package^35^ and the same or closely matched phenotypes as in LCV from opengwas.io^56^ to explore causes and consequences of PE. Independent genome-wide significant variants were selected as instruments, harmonized across exposure and outcome GWAS datasets, and analyzed using standard MR estimators (inverse-variance weighted, MR-Egger, and weighted median). We performed bidirectional MR using publicly available GWAS and maternal / fetal GWAS both as exposures and outcomes. We note that when using fetal GWAS as TwoSampleMR exposure, leaving us only two SNPs as exposure after filtering (rs4769612 and rs57741140, explaining ∼1.9% of PE variance), MR lacks power to detect effects below OR 1.33, with a criterion of *P*<0.00125 (corresponding for correction of 40 independent traits)^57^. Two-sample MR relies on independent instrument–exposure and instrument–outcome estimates, and shared individuals between the datasets can create unwanted correlation between the numerator and denominator of MR estimators^58^, which can be the case for some of the MR comparisons (Supplementary Tables 8&9)

## Supporting information

Non-author collaborators

## Data Availability

All data produced in the present study is either available in the supplements, or available upon a reasonable request to the authors. The source data utilized in this study can be accessed by applying to each of the participating studies (FINNPEC/FinnGen/Estonian Biobank/InterPregGen) individually.

## Acknowledgements

We sincerely thank all FINNPEC, FinnGen, Estonian Biobank and InterPregGen participants, principal investigators, laboratory personnel and data management teams.

The FinnGen project is funded by two grants from Business Finland (HUS 4685/31/2016 and UH 4386/31/2016) and the following industry partners: AbbVie Inc., AstraZeneca UK Ltd, Biogen MA Inc., Bristol Myers Squibb (and Celgene Corporation & Celgene International II Sàrl), Genentech Inc., Merck Sharp & Dohme LCC, Pfizer Inc., GlaxoSmithKline Intellectual Property Development Ltd., Sanofi US Services Inc., Maze Therapeutics Inc., Janssen Biotech Inc, Novartis AG, and Boehringer Ingelheim International GmbH. Following biobanks are acknowledged for delivering biobank samples to FinnGen: Auria Biobank (www.auria.fi/biopankki), THL Biobank (www.thl.fi/biobank), Helsinki Biobank (www.helsinginbiopankki.fi), Biobank Borealis of Northern Finland (https://www.ppshp.fi/Tutkimus-ja-opetus/Biopankki/Pages/Biobank-Borealis-briefly-in-English.aspx), Finnish Clinical Biobank Tampere (www.tays.fi/en-US/Research_and_development/Finnish_Clinical_Biobank_Tampere), Biobank of Eastern Finland (www.ita-suomenbiopankki.fi/en), Central Finland Biobank (www.ksshp.fi/fi-FI/Potilaalle/Biopankki), Finnish Red Cross Blood Service Biobank (www.veripalvelu.fi/verenluovutus/biopankkitoiminta), Terveystalo Biobank (www.terveystalo.com/fi/Yritystietoa/Terveystalo-Biopankki/Biopankki/) and Arctic Biobank (https://www.oulu.fi/en/university/faculties-and-units/faculty-medicine/northern-finland-birth-cohorts-and-arctic-biobank). All Finnish Biobanks are members of BBMRI.fi infrastructure (www.bbmri.fi). Finnish Biobank Cooperative - FINBB (https://finbb.fi/) is the coordinator of BBMRI-ERIC operations in Finland. The Finnish biobank data can be accessed through the Fingenious® services (https://site.fingenious.fi/en/) managed by FINBB.

This study was supported by Jane and Aatos Erkko Foundation (to H. Laivuori); the Päivikki and Sakari Sohlberg Foundation (A Kivelä, H. Laivuori); Academy of Finland (362074, 121196, 278941, 134957 to H. Laivuori); Research Funds of the University of Helsinki (to H. Laivuori); The Finnish Medical Foundation (to H. Laivuori); Finska Läkaresällskapet (to H. Laivuori); the Juho Vainio Foundation (to T. Jääskeläinen), EraPerMed JTC2020, Academy of Finland (344695, to H. Laivuori); the Competitive State Research Financing of the Expert Responsibility area of Helsinki University Hospital (to S. Heinonen), and Tampere University Hospital (to H. Laivuori). The Novo Nordisk Foundation, the Signe and Ane Gyllenberg Foundation, and the Foundation for Pediatric Research contributed to the FINNPEC (Finnish Genetics of Preeclampsia Consortium) Study.

## Ethical statements

The FINNPEC study was approved by the Coordinating Ethics Committee of the Hospital District of Helsinki and Uusimaa (149/EO/2007). All FINNPEC study participants and the participating parents of the neonates provided a written informed consent.

Study subjects in FinnGen provided informed consent for biobank research, based on the Finnish Biobank Act. Alternatively, separate research cohorts, collected prior the Finnish Biobank Act came into effect (in September 2013) and start of FinnGen (August 2017), were collected based on study-specific consents and later transferred to the Finnish biobanks after approval by Fimea (Finnish Medicines Agency), the National Supervisory Authority for Welfare and Health. Recruitment protocols followed the biobank protocols approved by Fimea. The Coordinating Ethics Committee of the Hospital District of Helsinki and Uusimaa (HUS) statement number for the FinnGen study is Nr HUS/990/2017.

The FinnGen study is approved by Finnish Institute for Health and Welfare (permit numbers: THL/2031/6.02.00/2017, THL/1101/5.05.00/2017, THL/341/6.02.00/2018, THL/2222/6.02.00/2018, THL/283/6.02.00/2019, THL/1721/5.05.00/2019 and THL/1524/5.05.00/2020), Digital and population data service agency (permit numbers: VRK43431/2017-3, VRK/6909/2018-3, VRK/4415/2019-3), the Social Insurance Institution (permit numbers: KELA 58/522/2017, KELA 131/522/2018, KELA 70/522/2019, KELA 98/522/2019, KELA 134/522/2019, KELA 138/522/2019, KELA 2/522/2020, KELA 16/522/2020), Findata permit numbers THL/2364/14.02/2020, THL/4055/14.06.00/2020, THL/3433/14.06.00/2020, THL/4432/14.06/2020, THL/5189/14.06/2020, THL/5894/14.06.00/2020, THL/6619/14.06.00/2020, THL/209/14.06.00/2021, THL/688/14.06.00/2021, THL/1284/14.06.00/2021, THL/1965/14.06.00/2021, THL/5546/14.02.00/2020, THL/2658/14.06.00/2021, THL/4235/14.06.00/2021, Statistics Finland (permit numbers: TK-53-1041-17 and TK/143/07.03.00/2020 (earlier TK-53-90-20) TK/1735/07.03.00/2021, TK/3112/07.03.00/2021) and Finnish Registry for Kidney Diseases permission/extract from the meeting minutes on 4^th^ July 2019.

The Biobank Access Decisions for FinnGen samples and data utilized in FinnGen Data Freeze 12 include: THL Biobank BB2017_55, BB2017_111, BB2018_19, BB_2018_34, BB_2018_67, BB2018_71, BB2019_7, BB2019_8, BB2019_26, BB2020_1, BB2021_65, Finnish Red Cross Blood Service Biobank 7.12.2017, Helsinki Biobank HUS/359/2017, HUS/248/2020, HUS/430/2021 §28, §29, HUS/150/2022 §12, §13, §14, §15, §16, §17, §18, §23, §58, §59, HUS/128/2023 §18, Auria Biobank AB17-5154 and amendment #1 (August 17 2020) and amendments BB_2021-0140, BB_2021-0156 (August 26 2021, Feb 2 2022), BB_2021-0169, BB_2021-0179, BB_2021-0161, AB20-5926 and amendment #1 (April 23 2020) and it’s modifications (Sep 22 2021), BB_2022-0262, BB_2022-0256, Biobank Borealis of Northern Finland_2017_1013, 2021_5010, 2021_5010 Amendment, 2021_5018, 2021_5018 Amendment, 2021_5015, 2021_5015 Amendment, 2021_5015 Amendment_2, 2021_5023, 2021_5023 Amendment, 2021_5023 Amendment_2, 2021_5017, 2021_5017 Amendment, 2022_6001, 2022_6001 Amendment, 2022_6006 Amendment, 2022_6006 Amendment, 2022_6006 Amendment_2, BB22-0067, 2022_0262, 2022_0262 Amendment, Biobank of Eastern Finland 1186/2018 and amendment 22§/2020, 53§/2021, 13§/2022, 14§/2022, 15§/2022, 27§/2022, 28§/2022, 29§/2022, 33§/2022, 35§/2022, 36§/2022, 37§/2022, 39§/2022, 7§/2023, 32§/2023, 33§/2023, 34§/2023, 35§/2023, 36§/2023, 37§/2023, 38§/2023, 39§/2023, 40§/2023, 41§/2023, Finnish Clinical Biobank Tampere MH0004 and amendments (21.02.2020 & 06.10.2020), BB2021-0140 8§/2021, 9§/2021, §9/2022, §10/2022, §12/2022, 13§/2022, §20/2022, §21/2022, §22/2022, §23/2022, 28§/2022, 29§/2022, 30§/2022, 31§/2022, 32§/2022, 38§/2022, 40§/2022, 42§/2022, 1§/2023, Central Finland Biobank 1-2017, BB_2021-0161, BB_2021-0169, BB_2021-0179, BB_2021-0170, BB_2022-0256, BB_2022-0262, BB22-0067, Decision allowing to continue data processing until 31^st^ Aug 2024 for projects: BB_2021-0179, BB22-0067,BB_2022-0262, BB_2021-0170, BB_2021-0164, BB_2021-0161, and BB_2021- 0169, and Terveystalo Biobank STB 2018001 and amendment 25^th^ Aug 2020, Finnish Hematological Registry and Clinical Biobank decision 18^th^ June 2021, Arctic biobank P0844: ARC_2021_1001.

The Estonian Biobank (EstBB) is a population-based biobank that has obtained clinical data from the national registries and hospital databases. Analyses in the EstBB were carried out under ethical approval 1.1-12/624 from the Estonian Committee on Bioethics, and Human Research and data release N05 from the EstBB. All biobank participants have signed a broad informed consent form.

