## Supplementary Figures for "Disentangling Maternal and Fetal Genetic Contributions to Preeclampsia"

**Supplementary Fig. 1. MAGMA tissue-specific gene expression enrichment for preeclampsia-associated Loci.**

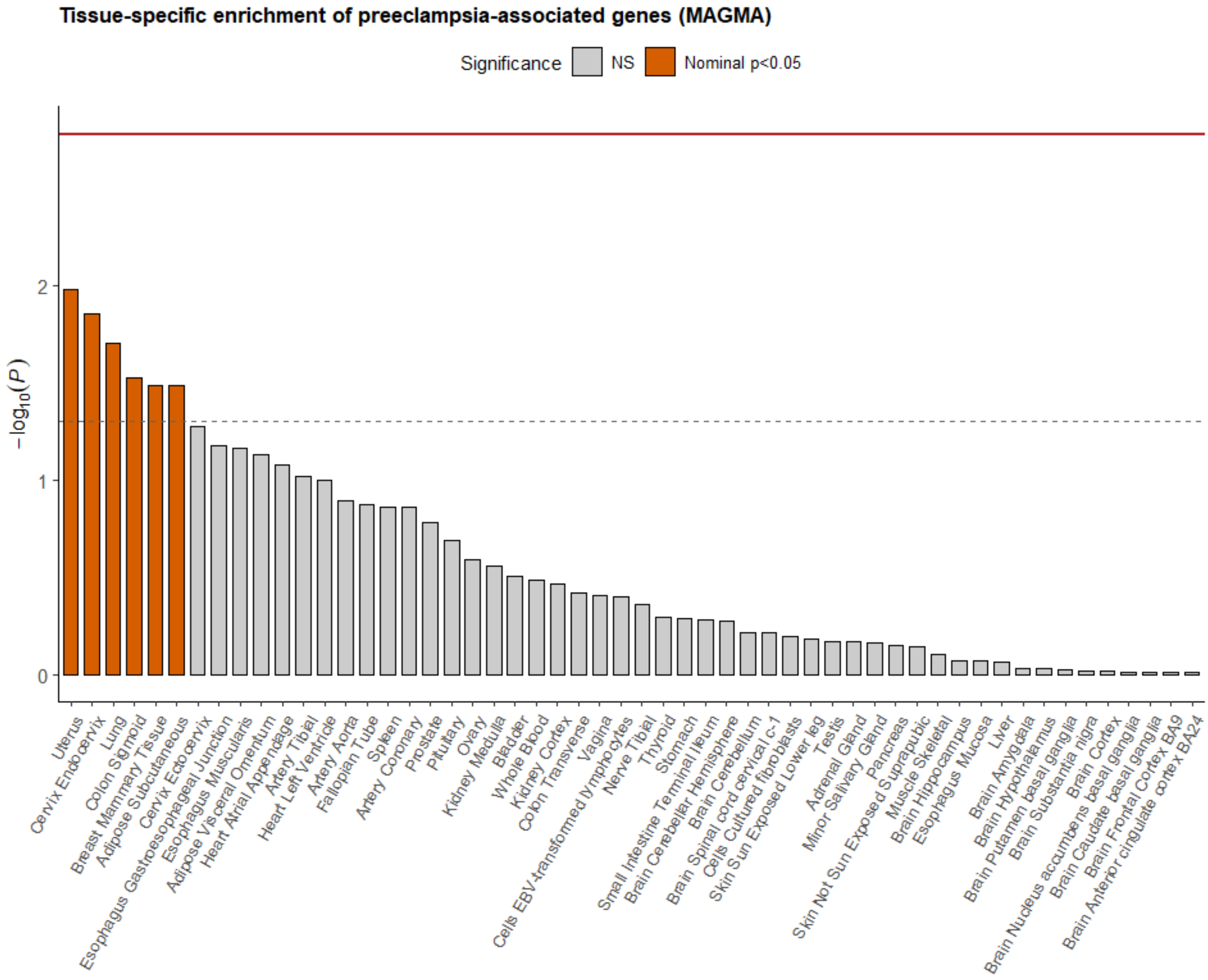


Tissue enrichment analysis was conducted using MAGMA implemented in the FUMA platform, testing whether genes associated with maternal preeclampsia show higher expression in 54 specific tissues based on GTEx v8 data. The x-axis indicates different tissue types and the y-axis shows the $-\log_{10}$-transformed *P*-values for enrichment. Orange bars indicate nominally significant enrichment to uterus (*P*=0.0105), endocervix (*P*=0.0139), lung (*P*=0.0198), colon sigmoid (*P*=0.0297), breast (*P*=0.0326), and subcutaneous adipose tissue (*P*=0.0328), suggesting biological relevance of these tissues to disease pathophysiology. Full results are available in Supplementary Table 5. None of the signals survived conservative Bonferroni correction for 30 general tissue types (red line).

**Supplementary Fig. 2. Manhattan plot of fetal effects on PE, highlighting loci below significance threshold of 1.0 x $10^{-6}$.**

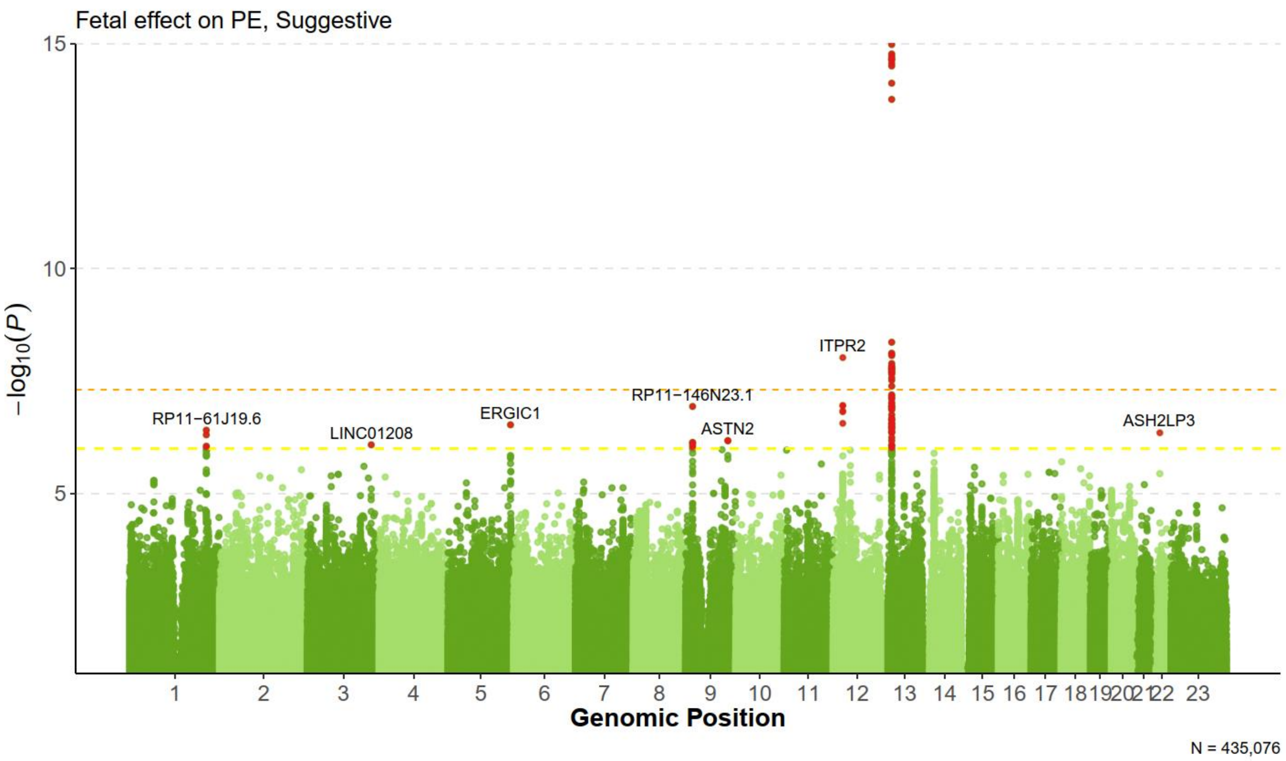


A Manhattan plot for fetal GWAS results, highlighting genes in 6 loci that contained variants with a *P*-value below 1.0 x$10^{6}$) (red dots), on top of the *EIF4A1P7/FLT1* signal in chromosome 13 and *ITPR2* locus in chromosome 12 (Supplementary Table 7). Yellow horizontal dashed line indicates the less stringent P-value, and orange horizontal dashed line the common GWAS threshold of 5.0 x10-[8.]

**Supplementary Fig. 3. Manhattan plot of multivariate GWAS on PE.**

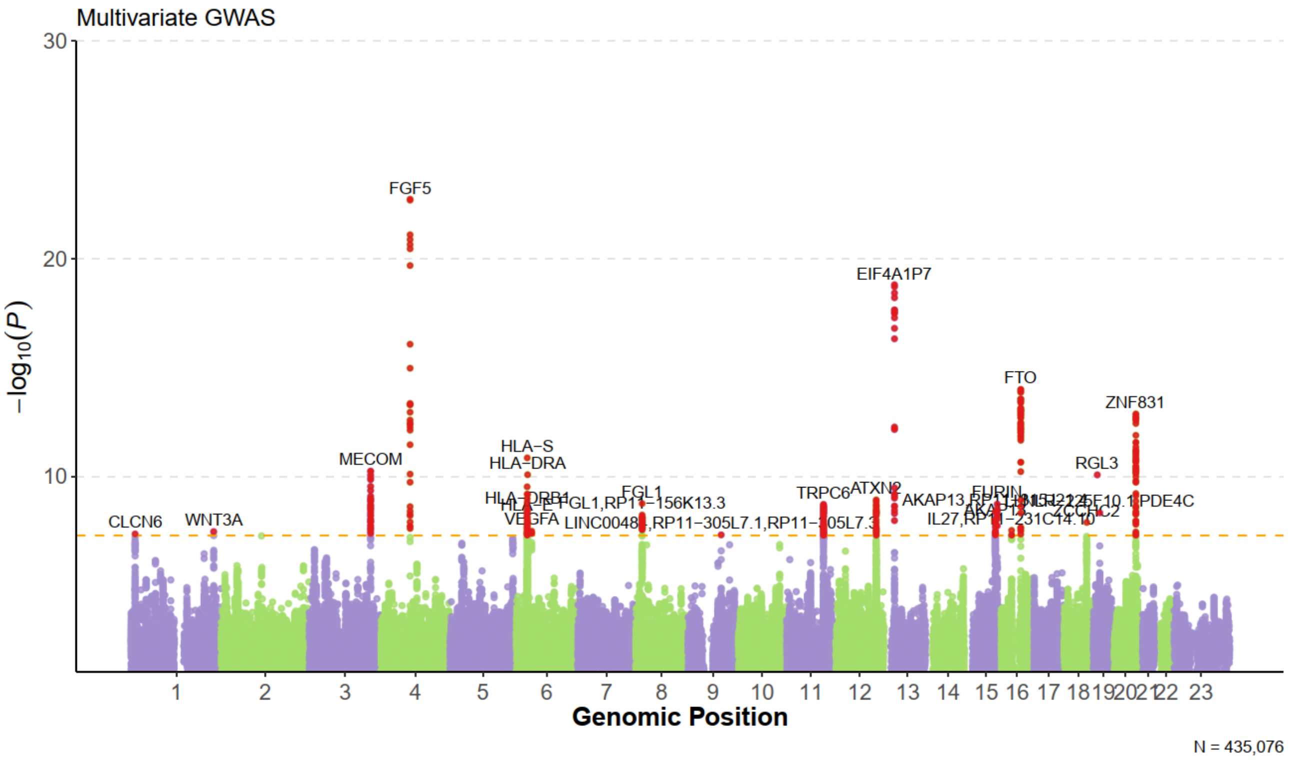


A Manhattan plot showing associations with PE based on multivariate GWAS analysis combining both maternal and fetal GWAS, based on 21,784 maternal cases and 379,813 maternal controls, and 8406 fetal cases and 426,670 fetal controls. The plot indicates the −log10-transformed *P* value of each tested variant along the vertical axis and their chromosomal positions along the horizontal axis. The orange line signifies the genome-wide significance threshold of *P* = 5.0 × 10−8. The genome-wide significant variants are indicated by red dots, with the nearest gene for each lead variant annotated. The analysis identified two additional genome-wide significant loci with lead variants, rs708119, close to *WNT3A* in chromosome 1 and rs57741140, close to *ZCCHC2* in chromosome 18.

**Supplementary Fig.4. Power analysis for paternal effects on PE**

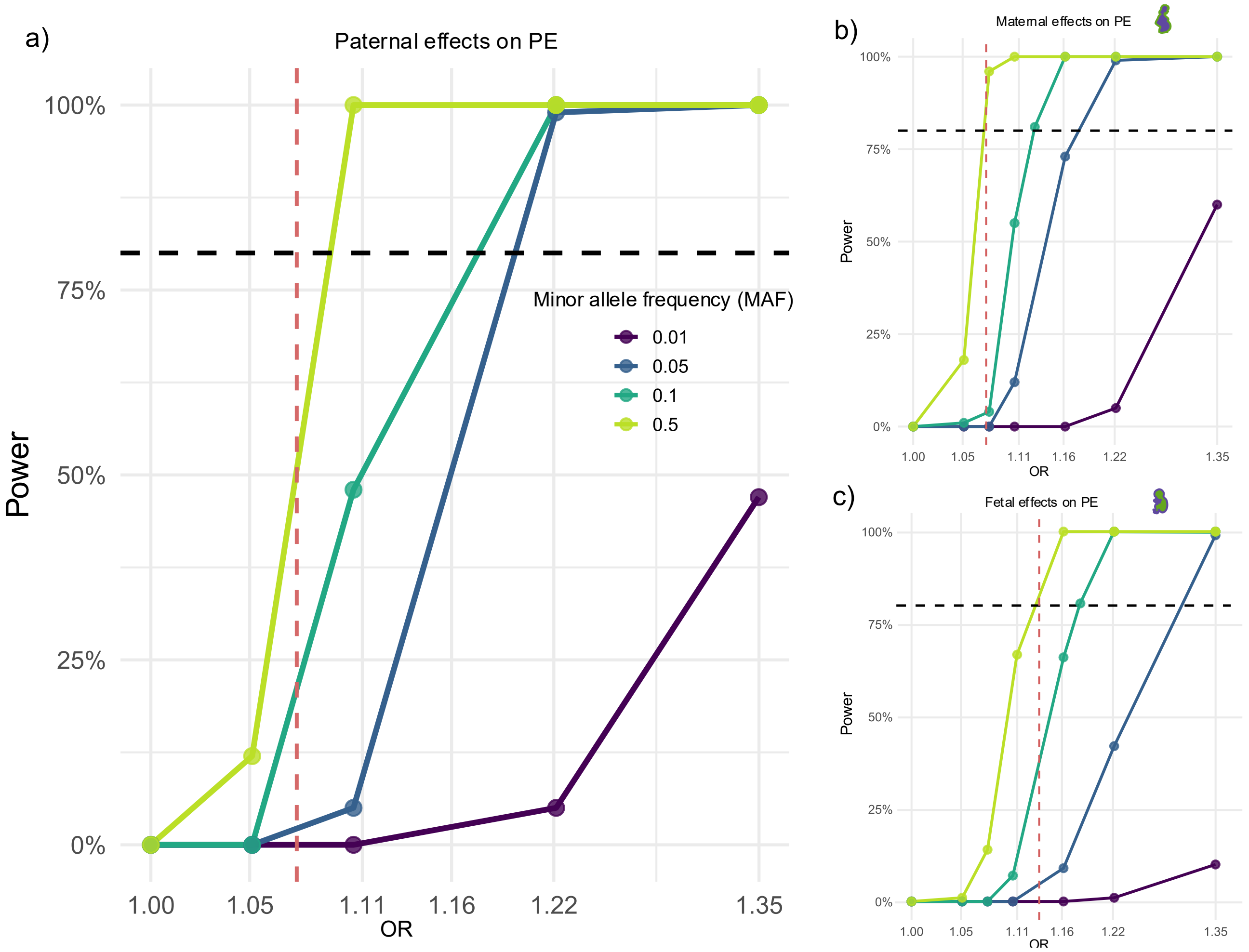


The paternal power curves (a) are based on simulations of 401,597 genotyped trios (mother, father, child) including 21,784 PE cases, based on the numbers from the maternal GWAS. For comparison, the plots illustrate also maternal (b) and fetal (c) power curves to associate variants at genome-wide significance threshold (*P*<5e-08) with PE. Like the paternal data, the estimated maternal power is based on simulations of 401,597 samples and 21,784 PE cases. The fetal power curves are based on simulations of 435,076 samples and 8406 cases. Light green = MAF 0.5. Green = MAF 0.1. Blue = MAF 0.05. Dark purple = MAF 0.01. Yellow bars = distribution of observed effect sizes for lead SNPs. Black horizontal dashed line = 80% power threshold. Red vertical dashed line = smallest observed effect size for a lead SNP in the maternal (a&b) and fetal (c) analyses. MAF=minor allele frequency, PE=preeclampsia.

### Supplementary Fig. 5. Genetic correlations and causality, WLM-corrected datasets

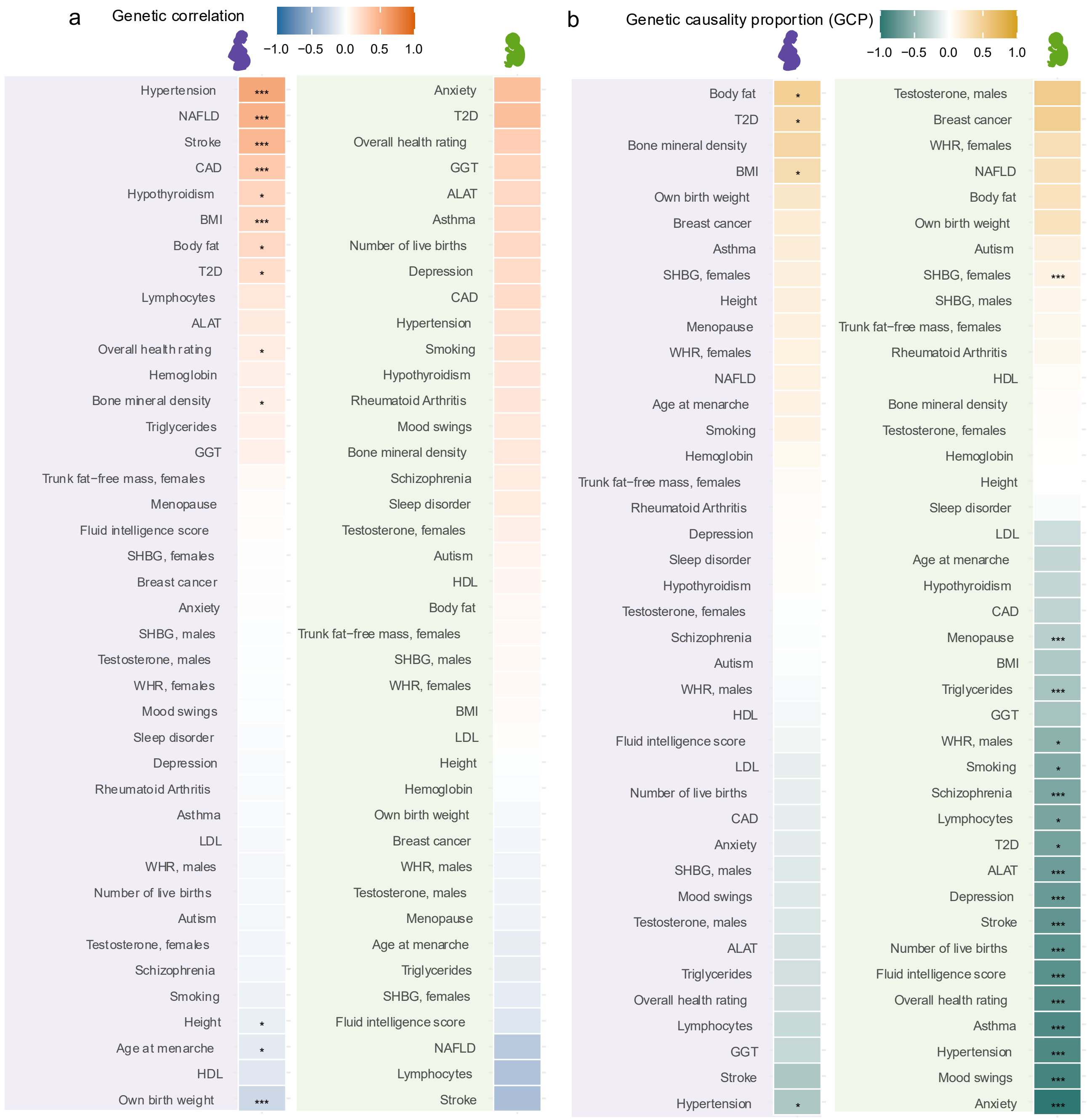


Genetic correlations and causality between the WLM-corrected maternal and fetal GWASs of preeclampsia (PE) and selected traits. The panel (a) shows genetic correlations, and panel (b) inferred causality from latent causal variable (LCV) analyses. Purple shadowing = results for WLM-corrected maternal genome wide association study (GWAS). Green shadowing = results for WLM-corrected fetal GWAS. Blue=negative genetic correlation, red=positive genetic correlation. Yellow = Causality of named traits on PE, Dark green = Causality of PE on the named traits. * *P*<0.05, *** *P*<0.00125, corresponding for conservative Bonferroni correction of 40 independent traits.

### Supplementary Fig. 6. Results from IVW MR-analyses, causes of maternal and fetal PE

a)

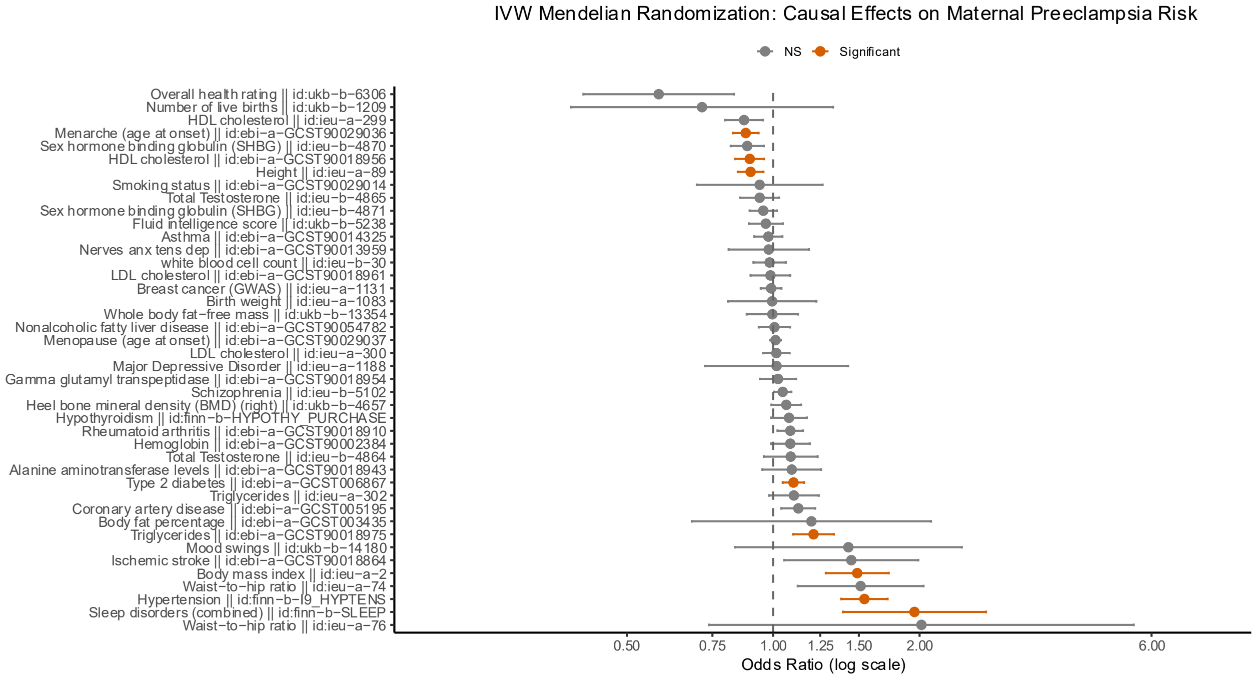


b)

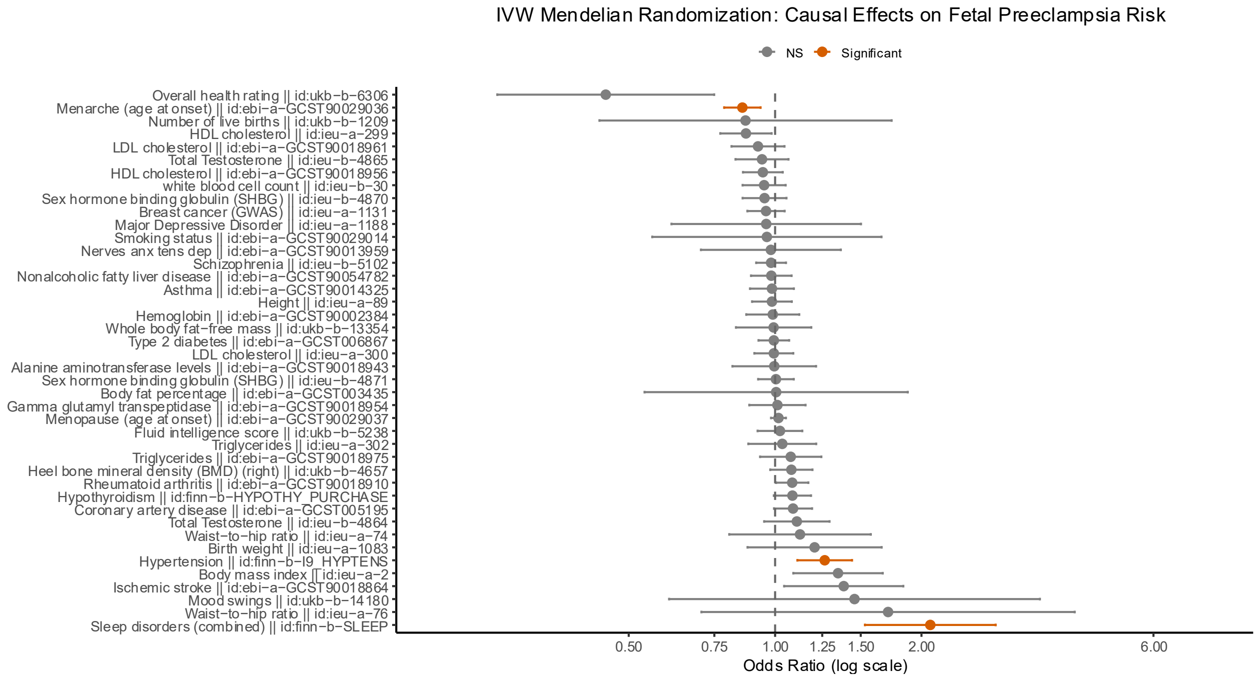


The figure summarizes results from IVW MR analyses using complex traits as exposures and maternal (a) and fetal (b) effects on PE as outcomes. Orange color indicates statistical significance after Bonferroni correction (*P*<0.00125). The analyses were performed using TwoSampleMR, using standard protocol and indicated phenotypes from opengwas.io.

### Supplementary Fig. 7. Results from IVW MR-analyses, causality of maternal and fetal PE

a)

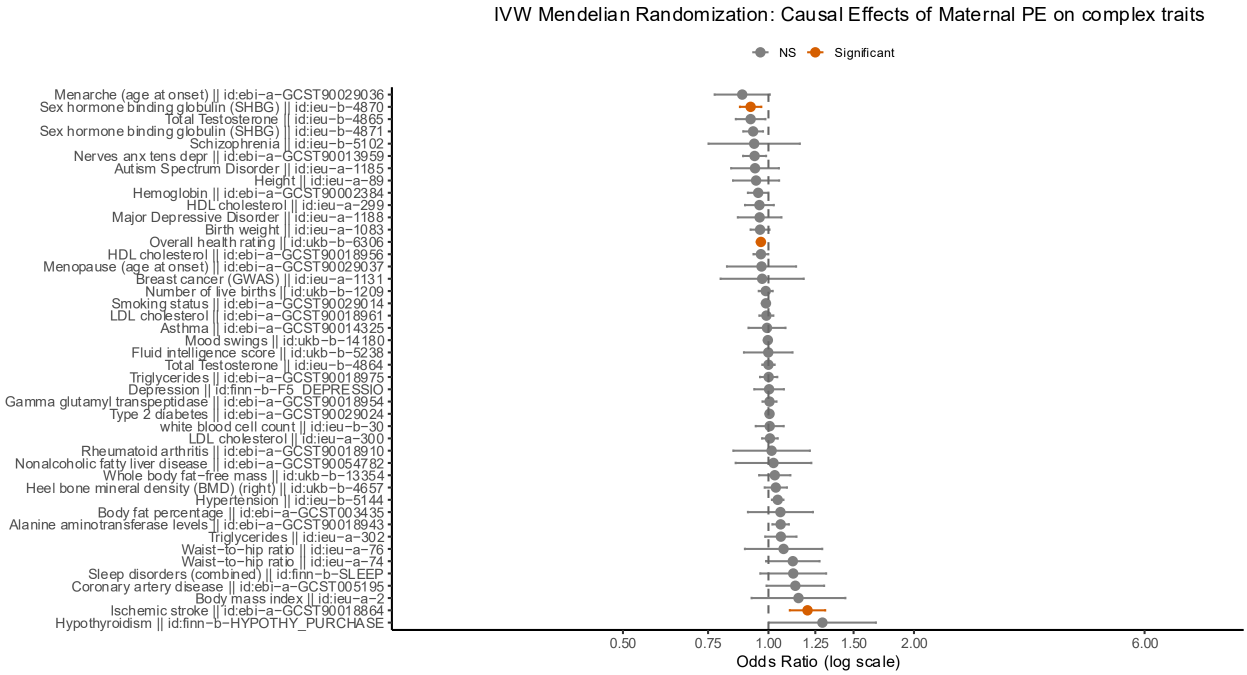


b)

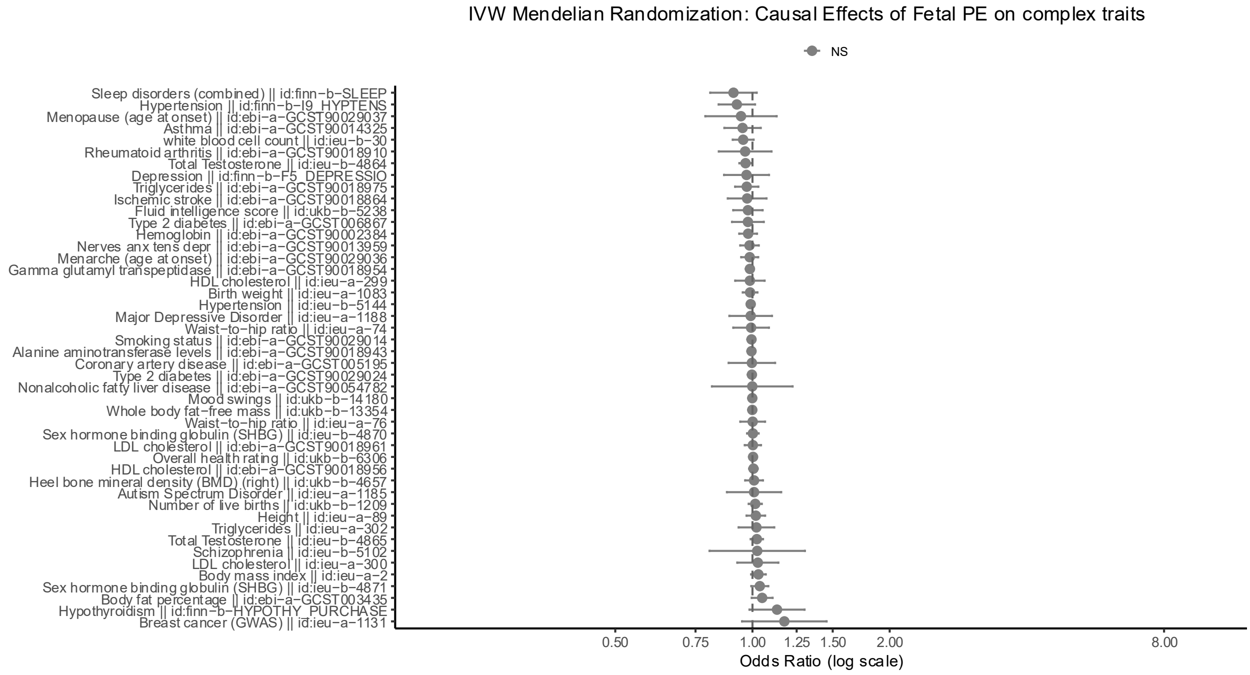


The figure summarizes results from IVW MR analyses using maternal (a) and fetal (b) effects on PE as exposures and complex traits as outcomes. Orange color indicates statistical significance after Bonferroni correction (*P*<0.00125). The analyses were performed using TwoSampleMR, using standard protocol and indicated phenotypes from opengwas.io.
