## Supplementary Code for "Disentangling Maternal and Fetal Genetic Contributions to Preeclampsia"

### **Estimating GWAS power in mother-child duos and mother-father-child trios for binary outcomes**

```
###########################################################################
#########SIMULATE POWER TO DETECT EFFECTS AT P<5e-08 IN MOTHER-CHILD DUOS###
###########################################################################

#######PLEASE NOTE THAT DEPENDING ON THE SIZE OF THE SIMULATED DATASET, THESE SIMULATIONS CAN BE
COMPUTATIONALLY HEAVY###############

sim_GWAS_power <- function(Nmc,Ncases,betao,betam,maf){

#########Create mother and child genotypes

# Generate allele frequencies under HWE
p_A <- 1 - maf  # Major allele frequency
p_a <- maf      # Minor allele frequency

# Sample one allele from a diploid genotype
sample_allele <- function(genotype) {
  if (genotype == 0) return(0)  # Homozygous major (AA)
  if (genotype == 2) return(1)  # Homozygous minor (aa)
  return(sample(c(0,1), size=1))  # Heterozygous (Aa) → Randomly pass major (0) or minor (1)
}

# Simulate child genotypes under HWE
child_genotype <- sample(c(0,1,2), size=Nmc, replace=TRUE,
                   prob=c(p_A^2, 2*p_A*p_a, p_a^2))

# Simulate the mother's alleles, ensuring 50% correlation
mother_genotype <- numeric(Nmc)

for (i in 1:Nmc) {
  child_allele_from_mother <- sample_allele(child_genotype[i])
  mother_alleles <- c(child_allele_from_mother, sample(c(0,1), size=1, prob=c(1-maf, maf)))
  mother_genotype[i] <- sum(mother_alleles)
}

genotype_matrix <- cbind(child_genotype, mother_genotype)
mc_var <- as.data.frame(genotype_matrix)

### Construct phenotype
### Set alpha and risk function

k = Ncases/Nmc
alpha = log(k/(1-k))
risk <- exp(alpha+betao*mc_var[,1]+betam*mc_var[,2])/(1+exp(alpha+betao*mc_var[,1]+betam*mc_var[,2]))
### Test power
# Case-control vector
cco <- rbinom(Nmc,size=1,risk)

########################
# Fit logistic regression model
########################

# Generalised linear regression: case status ~ offspring variant + maternal variant
out <- summary(glm(cco~mc_var[,1]+mc_var[,2],family='binomial'))$coefficients
# Output results: intercept, offspring beta, maternal beta, offspring p-value, maternal p-value
sim_res <- data.frame(alpha=out[1,1],betao=out[2,1],betam=out[3,1],pbetao=out[2,4],pbetam=out[3,4])
return(sim_res)
}
```

```
#####Example simulation for maternal effects on PE, in a dataset of 401000 samples and 21784 cases, with maternal
#####beta=0.05, fetal beta=0, MAF=0.01.
###########################################################################

# Run 100 iterations and store results
N_iter <- 100  # Number of simulations
results_list <- vector("list", N_iter)

for (i in 1:N_iter) {
  results_list[[i]] <- sim_GWAS_power(Nmc=401000, Ncases=21784, betao=0.0, betam=0.05, maf=0.01)
}

resbetam005_maf001 <- do.call(rbind, results_list) #### A result list that includes all simulations from 100 iterations

#####Finally, power is determined as the percentage of simulations with P<5e-08 (e.g. for 100 simulations 80% power equals
80 simulations with P<5e-08.

#####To create power curves, one needs to run several different simulations under desired settings.

########################################################################################
#########SIMULATE POWER TO DETECT EFFECTS AT P<5e-08 IN MOTHER-FATHER-CHILD TRIOS######
########################################################################################

sim_GWAS_power_trio <- function(Nmc, Ncases, betao, betam, betaf, maf) {

#######################
# Create genotypes for child, mother, and father
#######################

####Generate allele frequencies under HWE
p_A <- 1 - maf
p_a <- maf

# Sample one allele from a diploid genotype
sample_allele <- function(genotype) {
if (genotype == 0) return(0)  # Homozygous major (AA)
if (genotype == 2) return(1)  # Homozygous minor (aa)
return(sample(c(0,1), size = 1))  # Aa → randomly pass A or a
}

# Simulate paternal and maternal genotype under HWE
mother_genotype <- sample(c(0,1,2), size=Nmc, prob=c(p_A^2,2*p_A*p_a,p_a^2), replace=TRUE)
father_genotype <- sample(c(0,1,2), size=Nmc, prob=c(p_A^2,2*p_A*p_a,p_a^2), replace=TRUE)

# Simulate child's genotype (combining one allele from each parent)
child_genotype <- numeric(Nmc)
  for (i in 1:Nmc) {
    maternal_allele <- sample_allele(mother_genotype[i])
    paternal_allele <- sample_allele(father_genotype[i])
    child_genotype[i] <- maternal_allele + paternal_allele
  }

genotype_matrix <- data.frame(
child = child_genotype,
mother = mother_genotype,
father = father_genotype
)

### Construct phenotype
### Set alpha and risk function

k <- Ncases / Nmc
alpha <- log(k / (1 - k))
```

```
risk <- exp(alpha + betao * genotype_matrix$child + betam * genotype_matrix$mother + betaf * genotype_matrix$father) / (1 +
exp(alpha +betao * genotype_matrix$child + betam * genotype_matrix$mother +betaf * genotype_matrix$father))
# Generate binary case/control outcome
cco <- rbinom(Nmc, size = 1, prob = risk)

#######################
# Fit logistic regression model
#######################

model <- glm(cco ~ child + mother + father, data = genotype_matrix, family = "binomial")
out <- summary(model)$coefficients
#######################
# Output results
#######################
sim_res <- data.frame(alpha = out[1, 1],betao = out["child", 1],betam = out["mother", 1],betaf = out["father", 1],p_betao =
out["child", 4],p_betam = out["mother", 4],p_betaf = out["father", 4])
return(sim_res)
}

#######################
# Example simulation for paternal effects, with paternal beta 0.05, and maf 0.50, without maternal/fetal effects
#######################
N_iter <- 100
results_list <- vector("list", N_iter)
for (i in 1:N_iter) {
results_list[[i]] <- sim_GWAS_power_trio(
Nmc = 401000,
Ncases = 21784,
betao = 0.0,   # offspring effect
betam = 0.0,  # maternal effect
betaf = 0.05,  # paternal effect
maf = 0.50
)
}
# Combine into one dataframe
res_betaf005maf050 <- do.call(rbind, results_list)
```

#####Finally, power is determined as the percentage of simulations with P<5e-08 (e.g. for 100 simulations 80% power equals 80 simulations with P<5e-08.

#####To create power curves, one needs to run several different simulations under desired settings.
