## Supplementary material for "Disentangling Maternal and Fetal Genetic Contributions to Preeclampsia": Non-author collaborators

| Full Name | Study | Affiliation | Role 1 | Role 2 |
|---|---|---|---|---|
| Eeva Ekholm | FINNPEC | Department of Obstetrics and Gynecology, Turku University Hosp | Researcher in charge, Turku University Hospital | |
| Reija Hietala | FINNPEC | Department of Obstetrics and Gynecology, Helsinki University Ho | Researcher in charge, Helsinki University Hospital | |
| Leea Keski-Nisula | FINNPEC | Department of Obstetrics and Gynecology, University of Eastern | Researcher in charge, Kuopio University Hospital | |
| Kaarin Mäkikallio | FINNPEC | Department of Obstetrics and Gynecology, Turku University Hosp | Researcher in charge, Oulu University Hospital | |
| Jukka Uotila | FINNPEC | Department of Obstetrics and Gynecology, Tampere University He | Researcher in charge, Tampere University Hospital | |
| Susanna Sainio | FINNPEC | Finnish Red Cross Blood Service | Researcher in charge, Helsinki Maternity Hospital | |
| Terhi Saisto | FINNPEC | Department of Obstetrics and Gynecology, Helsinki University Ho | Researcher in charge, Helsinki University Hospital | |
| Marja Vääräsmäki | FINNPEC | Department of Obstetrics and Gynecology, Oulu University Hospit | Researcher in charge, Oulu University Hospital | |
| Tia Aalto-Viljakainen | FINNPEC | Oncology, Helsinki University Hospital and University of Helsinki | Clinical researcher | |
| Leena Georgiadis | FINNPEC | Obstetrics and Gynecology, Helsinki University Hospital. and Univ | Clinical researcher | |
| Jenni Heikkinen- Eloranta | FINNPEC | Obstetrics and Gynecology, Helsinki University Hospital. and Univ | Clinical researcher | |
| Miira M. Klemetti | FINNPEC | Obstetrics and Gynecology, Helsinki University Hospital. and Univ | Clinical researcher | |
| Sanna Suomalainen- König | FINNPEC | Obstetrics and Gynecology, Helsinki University Hospital. and Univ | Clinical researcher | |
| Satu Wedenoja | FINNPEC | Obstetrics and Gynecology, University of Helsinki | Clinical researcher | |
| Satu Leminen | FINNPEC | Obstetrics and Gynecology, Helsinki University Hospital and Unive | Research nurse | |
| Aija Lähdesmäki | FINNPEC | Finnish Institute for Health and Welfare | Research nurse | |
| Susanna Mehtälä | FINNPEC | Department of Obstetrics and Gynecology, Helsinki University Hos | Technician | |
| Christina Salmén | FINNPEC | Cardiology, Helsinki University Hospital | Research nurse | |
| Aarno Palotie | FinnGen | Institute for Molecular Medicine Finland (FIMM), HiLIFE, | Steering Committee | Steering Committee |
| Mark Daly | FinnGen | Institute for Molecular Medicine Finland (FIMM), HiLIFE, | Steering Committee | Steering Committee |
| Bridget Riley-Gills | FinnGen | Abbvie, Chicago, IL, United States | Steering Committee | Pharmaceutical companies |
| Howard Jacob | FinnGen | Abbvie, Chicago, IL, United States | Steering Committee | Pharmaceutical companies |
| Coralie Viollet | FinnGen | Astra Zeneca, Cambridge, United Kingdom | Steering Committee | Pharmaceutical companies |
| Slavé Petrovski | FinnGen | Astra Zeneca, Cambridge, United Kingdom | Steering Committee | Pharmaceutical companies |
| Alix Berton | FinnGen | Bayer AG, Leverkusen, Germany | Steering Committee | Pharmaceutical companies |
| Santha Ramakrishnan | FinnGen | Bayer AG, Leverkusen, Germany | Steering Committee | Pharmaceutical companies |
| Ellen Tsai | FinnGen | Biogen, Cambridge, MA, United States | Steering Committee | Pharmaceutical companies |
| Zhihao Ding | FinnGen | Boehringer Ingelheim, Ingelheim am Rhein, Germany | Steering Committee | Pharmaceutical companies |
| Emily Holzinger | FinnGen | Bristol Myers Squibb, New York, NY, United States | Steering Committee | Pharmaceutical companies |
| Robert Plenge | FinnGen | Bristol Myers Squibb, New York, NY, United States | Steering Committee | Pharmaceutical companies |
| Joseph Maranville | FinnGen | Bristol Myers Squibb, New York, NY, United States | Steering Committee | Pharmaceutical companies |
| Mark McCarthy | FinnGen | Genentech, San Francisco, CA, United States | Steering Committee | Pharmaceutical companies |
| Rion Pendergrass | FinnGen | Genentech, San Francisco, CA, United States | Steering Committee | Pharmaceutical companies |
| Jonathan Davitte | FinnGen | GlaxoSmithKline, Collegeville, PA, United States | Steering Committee | Pharmaceutical companies |
| Chia-Yen Chen | FinnGen | Merck, Kenilworth, NJ, United States | Steering Committee | Pharmaceutical companies |
| Melis Atalar Aksit | FinnGen | Pfizer, New York, NY, United States | Steering Committee | Pharmaceutical companies |
| Anna Vlahiotis | FinnGen | Pfizer, New York, NY, United States | Steering Committee | Pharmaceutical companies |
| Katherine Klinger | FinnGen | Translational Sciences, Sanofi R&D, Framingham, MA, USA | Steering Committee | Pharmaceutical companies |
| Clement Chatelain | FinnGen | Translational Sciences, Sanofi R&D, Framingham, MA, USA | Steering Committee | Pharmaceutical companies |
| Jorg Blankenstein | FinnGen | Translational Sciences, Sanofi R&D, Framingham, MA, USA | Steering Committee | Pharmaceutical companies |
| Karol Estrada | FinnGen | Maze Therapeutics, San Francisco, CA, United States | Steering Committee | Pharmaceutical companies |
| Robert Graham | FinnGen | Maze Therapeutics, San Francisco, CA, United States | Steering Committee | Pharmaceutical companies |
| Dawn Waterworth | FinnGen | Johnson & Johnson Innovative Medicine, Spring House, PA, | Steering Committee | Pharmaceutical companies |
| Chris O´Donnell | FinnGen | Novartis Institutes for BioMedical Research, Cambridge, MA, Unite | Steering Committee | Pharmaceutical companies |
| Nicole Renaud | FinnGen | Novartis Institutes for BioMedical Research, Cambridge, MA, Unite | Steering Committee | Pharmaceutical companies |
| Tomi P. Mäkelä | FinnGen | HiLIFE, University of Helsinki, Finland, Finland | Steering Committee | University of Helsinki & Biobanks |
| Jaakko Kaprio | FinnGen | Institute for Molecular Medicine Finland (FIMM), HiLIFE, | Steering Committee | University of Helsinki & Biobanks |
| Minna Ruddock | FinnGen | Arctic biobank / University of Oulu | Steering Committee | University of Helsinki & Biobanks |
| Lila Kallio | FinnGen | Auria Biobank / University of Turku / Wellbeing Services County of | Steering Committee | University of Helsinki & Biobanks |
| Antti Hakanen | FinnGen | Auria Biobank / University of Turku / Wellbeing Services County of | Steering Committee | University of Helsinki & Biobanks |
| Terhi Kilpi | FinnGen | THL Biobank / Finnish Institute for Health and Welfare (THL), | Steering Committee | University of Helsinki & Biobanks |
| Markus Perola | FinnGen | THL Biobank / Finnish Institute for Health and Welfare (THL), | Steering Committee | University of Helsinki & Biobanks |
| Jukka Partanen | FinnGen | Finnish Red Cross Blood Service / Finnish Hematology Registry ar | Steering Committee | University of Helsinki & Biobanks |
| Taneli Raivio | FinnGen | Helsinki Biobank / Helsinki University and Hospital District of | Steering Committee | University of Helsinki & Biobanks |
| Eero Punkka | FinnGen | Helsinki Biobank / Helsinki University and Hospital District of | Steering Committee | University of Helsinki & Biobanks |
| Teija Kekonen | FinnGen | Northern Finland Biobank Borealis / University of Oulu / Wellbeing | Steering Committee | University of Helsinki & Biobanks |
| Raisa Serpi | FinnGen | Northern Finland Biobank Borealis / University of Oulu / Wellbeing | Steering Committee | University of Helsinki & Biobanks |
| Kati Kristiansson | FinnGen | Finnish Clinical Biobank Tampere / University of Tampere / | Steering Committee | University of Helsinki & Biobanks |
| Sanna Siltanen | FinnGen | Finnish Clinical Biobank Tampere / University of Tampere / | Steering Committee | University of Helsinki & Biobanks |
| Veli-Matti Kosma | FinnGen | Biobank of Eastern Finland / University of Eastern Finland / | Steering Committee | University of Helsinki & Biobanks |
| Arto Mannermaa | FinnGen | Biobank of Eastern Finland / University of Eastern Finland / | Steering Committee | University of Helsinki & Biobanks |
| Jari Laukkanen | FinnGen | Central Finland Biobank / University of Jyväskylä / Wellbeing | Steering Committee | University of Helsinki & Biobanks |
| Tiina Jokela | FinnGen | Central Finland Biobank / University of Jyväskylä / Wellbeing | Steering Committee | University of Helsinki & Biobanks |
| Mervi Ahlroth | FinnGen | Finnish Biobank Cooperative - FINBB | Steering Committee | University of Helsinki & Biobanks |
| Johanna Mäkelä | FinnGen | Finnish Biobank Cooperative - FINBB | Steering Committee | University of Helsinki & Biobanks |
| Outi Tuovila | FinnGen | Business Finland, Helsinki, Finland | Steering Committee | Other Experts/ Non-Voting Members |
| Jeffrey Waring | FinnGen | Abbvie, Chicago, IL, United States | Scientific Committee | Pharmaceutical companies |
| Bridget Riley-Gillis | FinnGen | Abbvie, Chicago, IL, United States | Scientific Committee | Pharmaceutical companies |
| Fedik Rahimov | FinnGen | Abbvie, Chicago, IL, United States | Scientific Committee | Pharmaceutical companies |
| Ioanna Tachmazidou | FinnGen | Astra Zeneca, Cambridge, United Kingdom | Scientific Committee | Pharmaceutical companies |
| Slavé Petrovski | FinnGen | Astra Zeneca, Cambridge, United Kingdom | Scientific Committee | Pharmaceutical companies |
| Alix Berton | FinnGen | Bayer AG, Leverkusen, Germany | Scientific Committee | Pharmaceutical companies |
| Santha Ramakrishnan | FinnGen | Bayer AG, Leverkusen, Germany | Scientific Committee | Pharmaceutical companies |
| Ellen Tsai | FinnGen | Biogen, Cambridge, MA, United States | Scientific Committee | Pharmaceutical companies |
| Zhihao Ding | FinnGen | Boehringer Ingelheim, Ingelheim am Rhein, Germany | Scientific Committee | Pharmaceutical companies |
| Marc Jung | FinnGen | Boehringer Ingelheim, Ingelheim am Rhein, Germany | Scientific Committee | Pharmaceutical companies |
| Hanati Tuoken | FinnGen | Boehringer Ingelheim, Ingelheim am Rhein, Germany | Scientific Committee | Pharmaceutical companies |
| Shameek Biswas | FinnGen | Bristol Myers Squibb, New York, NY, United States | Scientific Committee | Pharmaceutical companies |
| Benjamin Sun | FinnGen | Bristol Myers Squibb, New York, NY, United States | Scientific Committee | Pharmaceutical companies |
| Rion Pendergrass | FinnGen | Genentech, San Francisco, CA, United States | Scientific Committee | Pharmaceutical companies |
| Jonathan Davitte | FinnGen | GlaxoSmithKline, Collegeville, PA, United States | Scientific Committee | Pharmaceutical companies |
| Neha Raghavan | FinnGen | Merck, Kenilworth, NJ, United States | Scientific Committee | Pharmaceutical companies |
| Jae-Hoon Sul | FinnGen | Merck, Kenilworth, NJ, United States | Scientific Committee | Pharmaceutical companies |
| Melis Atalar Aksit | FinnGen | Pfizer, New York, NY, United States | Scientific Committee | Pharmaceutical companies |
| Xinli Hu | FinnGen | Pfizer, New York, NY, United States | Scientific Committee | Pharmaceutical companies |
| Katherine Klinger | FinnGen | Translational Sciences, Sanofi R&D, Framingham, MA, USA | Scientific Committee | Pharmaceutical companies |
| Robert Graham | FinnGen | Maze Therapeutics, San Francisco, CA, United States | Scientific Committee | Pharmaceutical companies |
| Dawn Waterworth | FinnGen | Johnson & Johnson Innovative Medicine, Spring House, PA, | Scientific Committee | Pharmaceutical companies |
| Nicole Renaud | FinnGen | Novartis Institutes for BioMedical Research, Cambridge, MA, Unite | Scientific Committee | Pharmaceutical companies |
| Ma´en Obeidat | FinnGen | Novartis Institutes for BioMedical Research, Cambridge, MA, Unite | Scientific Committee | Pharmaceutical companies |
| Jonathan Chung | FinnGen | Novartis Institutes for BioMedical Research, Cambridge, MA, Unite | Scientific Committee | Pharmaceutical companies |
| Jonas Zierer | FinnGen | Novartis Institutes for BioMedical Research, Cambridge, MA, Unite | Scientific Committee | Pharmaceutical companies |
| Mari Niemi | FinnGen | Novartis Institutes for BioMedical Research, Cambridge, MA, Unite | Scientific Committee | Pharmaceutical companies |
| Samuli Ripatti | FinnGen | Institute for Molecular Medicine Finland (FIMM), HiLIFE, University | Scientific Committee | University of Helsinki & Biobanks |

| | | | | |
|---|---|---|---|---|
| Johanna Schleutker | FinnGen | Auria Biobank / University of Turku / Wellbeing Services County of | Scientific Committee | University of Helsinki & Biobanks |
| Markus Perola | FinnGen | THL Biobank / Finnish Institute for Health and Welfare (THL), Hels | Scientific Committee | University of Helsinki & Biobanks |
| Tiina Wahlfors | FinnGen | THL Biobank / Finnish Institute for Health and Welfare (THL), Hels | Scientific Committee | University of Helsinki & Biobanks |
| Mikko Arvas | FinnGen | Finnish Red Cross Blood Service / Finnish Hematology Registry ar | Scientific Committee | University of Helsinki & Biobanks |
| Olli Carpén | FinnGen | Helsinki Biobank / Helsinki University and Hospital District of | Scientific Committee | University of Helsinki & Biobanks |
| Reetta Hinttala | FinnGen | Northern Finland Biobank Borealis / University of Oulu / Wellbeing | Scientific Committee | University of Helsinki & Biobanks |
| Johannes Kettunen | FinnGen | Northern Finland Biobank Borealis / University of Oulu / Wellbeing | Scientific Committee | University of Helsinki & Biobanks |
| Arto Mannermaa | FinnGen | Biobank of Eastern Finland / University of Eastern Finland / | Scientific Committee | University of Helsinki & Biobanks |
| Katriina Aalto-Setälä | FinnGen | Faculty of Medicine and Health Technology, Tampere University, T | Scientific Committee | University of Helsinki & Biobanks |
| Mika Kähönen | FinnGen | Finnish Clinical Biobank Tampere / University of Tampere / | Scientific Committee | University of Helsinki & Biobanks |
| Jari Laukkanen | FinnGen | Central Finland Biobank / University of Jyväskylä / Wellbeing | Scientific Committee | University of Helsinki & Biobanks |
| Johanna Mäkelä | FinnGen | FINBB - Finnish biobank cooperative | Scientific Committee | University of Helsinki & Biobanks |
| Hanna Kujala | FinnGen | Biobank of Eastern Finland / University of Eastern Finland / | Clinical Group / Task Force | |
| Triin Laisk | FinnGen | Estonian biobank, Tartu, Estonia | Clinical Group / Task Force | |
| Natalia Pujol | FinnGen | Estonian biobank, Tartu, Estonia | Clinical Group / Task Force | |
| Mika Kähönen | FinnGen | Finnish Clinical Biobank Tampere / University of Tampere / | Clinical Group / Task Force | |
| Veikko Salomaa | FinnGen | Finnish Institute for Health and Welfare (THL), Helsinki, Finland | Clinical Group / Task Force | |
| Jaana Suvisaari | FinnGen | Finnish Institute for Health and Welfare (THL), Helsinki, Finland | Clinical Group / Task Force | |
| Satu Koskela | FinnGen | Finnish Red Cross Blood Service / Finnish Hematology Registry ar | Clinical Group / Task Force | |
| Jouni Lauronen | FinnGen | Finnish Red Cross Blood Service / Finnish Hematology Registry ar | Clinical Group / Task Force | |
| Kristiina Aittomäki | FinnGen | Helsinki University Central Hospital, Helsinki, Finland | Clinical Group / Task Force | |
| Pirkko Pussinen | FinnGen | Helsinki University Hospital and University of Helsinki, Helsinki / | Clinical Group / Task Force | |
| Tuomo Meretoja | FinnGen | Helsinki University Hospital and University of Helsinki, Helsinki, Fir | Clinical Group / Task Force | |
| Heikki Joensuu | FinnGen | Helsinki University Hospital and University of Helsinki, Helsinki, Fir | Clinical Group / Task Force | |
| Peeter Karihtala | FinnGen | Helsinki University Hospital and University of Helsinki, Helsinki, Fir | Clinical Group / Task Force | |
| Emma Juuri | FinnGen | Helsinki University Hospital and University of Helsinki, Helsinki, Fir | Clinical Group / Task Force | |
| Aino Salminen | FinnGen | Helsinki University Hospital and University of Helsinki, Helsinki, Fir | Clinical Group / Task Force | |
| Tuula Salo | FinnGen | Helsinki University Hospital and University of Helsinki, Helsinki, Fir | Clinical Group / Task Force | |
| David Rice | FinnGen | Helsinki University Hospital and University of Helsinki, Helsinki, Fir | Clinical Group / Task Force | |
| Pekka Nieminen | FinnGen | Helsinki University Hospital and University of Helsinki, Helsinki, Fir | Clinical Group / Task Force | |
| Ulla Palotie | FinnGen | Helsinki University Hospital and University of Helsinki, Helsinki, Fir | Clinical Group / Task Force | |
| Fredrik Åberg | FinnGen | Helsinki University Hospital and University of Helsinki, Helsinki, Fir | Clinical Group / Task Force | |
| Daniel Gordin | FinnGen | Helsinki University Hospital and University of Helsinki, Helsinki, Fir | Clinical Group / Task Force | |
| Patrik Finne | FinnGen | Helsinki University Hospital and University of Helsinki, Helsinki, Fir | Clinical Group / Task Force | |
| Joni A Turunen | FinnGen | Helsinki University Hospital and University of Helsinki, Helsinki, Fir | Clinical Group / Task Force | |
| Minna Raivio | FinnGen | Hospital District of Helsinki and Uusimaa, Helsinki, Finland | Clinical Group / Task Force | |
| Pentti Tienari | FinnGen | Hospital District of Helsinki and Uusimaa, Helsinki, Finland | Clinical Group / Task Force | |
| Martti Färkkilä | FinnGen | Hospital District of Helsinki and Uusimaa, Helsinki, Finland | Clinical Group / Task Force | |
| Jukka Koskela | FinnGen | Hospital District of Helsinki and Uusimaa, Helsinki, Finland | Clinical Group / Task Force | |
| Sampsa Pikkarainen | FinnGen | Hospital District of Helsinki and Uusimaa, Helsinki, Finland | Clinical Group / Task Force | |
| Kari Eklund | FinnGen | Hospital District of Helsinki and Uusimaa, Helsinki, Finland | Clinical Group / Task Force | |
| Paula Kauppi | FinnGen | Hospital District of Helsinki and Uusimaa, Helsinki, Finland | Clinical Group / Task Force | |
| Daniel Gordin | FinnGen | Hospital District of Helsinki and Uusimaa, Helsinki, Finland | Clinical Group / Task Force | |
| Juha Sinisalo | FinnGen | Hospital District of Helsinki and Uusimaa, Helsinki, Finland | Clinical Group / Task Force | |
| Marja-Riitta Taskinen | FinnGen | Hospital District of Helsinki and Uusimaa, Helsinki, Finland | Clinical Group / Task Force | |
| Tiinamaija Tuomi | FinnGen | Hospital District of Helsinki and Uusimaa, Helsinki, Finland | Clinical Group / Task Force | |
| Timo Hiltunen | FinnGen | Hospital District of Helsinki and Uusimaa, Helsinki, Finland | Clinical Group / Task Force | |
| Johanna Mattson | FinnGen | Hospital District of Helsinki and Uusimaa, Helsinki, Finland | Clinical Group / Task Force | |
| Eveliina Salminen | FinnGen | Hospital District of Helsinki and Uusimaa, Helsinki, Finland | Clinical Group / Task Force | |
| Terhi Ollila | FinnGen | Hospital District of Helsinki and Uusimaa, Helsinki, Finland | Clinical Group / Task Force | |
| Katariina Hannula-Jouppi | FinnGen | Hospital District of Helsinki and Uusimaa, Helsinki, Finland | Clinical Group / Task Force | |
| Oskari Heikinheimo | FinnGen | Hospital District of Helsinki and Uusimaa, Helsinki, Finland | Clinical Group / Task Force | |
| Ilkka Kalliala | FinnGen | Hospital District of Helsinki and Uusimaa, Helsinki, Finland | Clinical Group / Task Force | |
| Lauri Aaltonen | FinnGen | Hospital District of Helsinki and Uusimaa, Helsinki, Finland | Clinical Group / Task Force | |
| Erkki Isometsä | FinnGen | Hospital District of Helsinki and Uusimaa, Helsinki, Finland | Clinical Group / Task Force | |
| Antti Aarnisalo | FinnGen | Hospital District of Helsinki and Uusimaa, Helsinki, Finland | Clinical Group / Task Force | |
| Ilkka Immonen | FinnGen | Hospital District of Helsinki and Uusimaa, Helsinki, Finland | Clinical Group / Task Force | |
| Salla Ranta | FinnGen | Hospital District of Helsinki and Uusimaa, Helsinki, Finland | Clinical Group / Task Force | |
| Filip Scheperjans | FinnGen | Hospital District of Helsinki and Uusimaa, Helsinki, Finland | Clinical Group / Task Force | |
| Felix Vaura | FinnGen | Institute for Molecular Medicine Finland (FIMM), HiLIFE, University | Clinical Group / Task Force | |
| Nina Mars | FinnGen | Institute for Molecular Medicine Finland (FIMM), HiLIFE, University | Clinical Group / Task Force | |
| Esa Pitkänen | FinnGen | Institute for Molecular Medicine Finland (FIMM), HiLIFE, University | Clinical Group / Task Force | |
| Hannele Laivuori | FinnGen | Institute for Molecular Medicine Finland (FIMM), HiLIFE, University | Clinical Group / Task Force | |
| Katja Kivinen | FinnGen | Institute for Molecular Medicine Finland (FIMM), HiLIFE, University | Clinical Group / Task Force | |
| Elisabeth Widen | FinnGen | Institute for Molecular Medicine Finland (FIMM), HiLIFE, University | Clinical Group / Task Force | |
| Taru Tukiainen | FinnGen | Institute for Molecular Medicine Finland (FIMM), HiLIFE, University | Clinical Group / Task Force | |
| Hanna Ollila | FinnGen | Institute for Molecular Medicine Finland (FIMM), HiLIFE, University | Clinical Group / Task Force | |
| Elmo Saarentaus | FinnGen | Institute for Molecular Medicine Finland (FIMM), HiLIFE, University | Clinical Group / Task Force | |
| Anne Kerola | FinnGen | Institute for Molecular Medicine Finland (FIMM), HiLIFE, University | Clinical Group / Task Force | |
| Eero Vuoksimaa | FinnGen | Institute for Molecular Medicine Finland (FIMM), HiLIFE, University | Clinical Group / Task Force | |
| Joni Lindbohm | FinnGen | Institute for Molecular Medicine Finland (FIMM), HiLIFE, University | Clinical Group / Task Force | |
| Zhiyu Yang | FinnGen | Institute for Molecular Medicine Finland (FIMM), HiLIFE, University | Clinical Group / Task Force | |
| Matthew Sampson | FinnGen | Institute for Molecular Medicine Finland (FIMM), HiLIFE, | Clinical Group / Task Force | |
| Adrian Banerji | FinnGen | Institute for Molecular Medicine Finland (FIMM), HiLIFE, | Clinical Group / Task Force | |
| Michelle McNulty | FinnGen | Institute for Molecular Medicine Finland (FIMM), HiLIFE, | Clinical Group / Task Force | |
| Aoxing Liu | FinnGen | Institute for Molecular Medicine Finland (FIMM), HiLIFE, | Clinical Group / Task Force | |
| Joel Rämö | FinnGen | Institute for Molecular Medicine Finland (FIMM), HiLIFE, | Clinical Group / Task Force | |
| Austin Argentieri | FinnGen | Institute for Molecular Medicine Finland (FIMM), HiLIFE, | Clinical Group / Task Force | |
| Amanda Elliott | FinnGen | Institute for Molecular Medicine Finland (FIMM), HiLIFE, | Clinical Group / Task Force | |
| Elisa Rahikkala | FinnGen | Northern Ostrobothnia Hospital District, Oulu, Finland | Clinical Group / Task Force | |
| Kirsi Sipilä | FinnGen | Oulu University Hospital and University of Oulu, Oulu, Finland | Clinical Group / Task Force | |
| Valtteri Julkunen | FinnGen | University of Eastern Finland and Kuopio University Hospital, Kuop | Clinical Group / Task Force | |
| Ville Leinonen | FinnGen | University of Eastern Finland and Kuopio University Hospital, Kuop | Clinical Group / Task Force | |
| Sanna Toppila-Salmi | FinnGen | University of Eastern Finland and Kuopio University Hospital, | Clinical Group / Task Force | |
| Mikko Hiltunen | FinnGen | University of Eastern Finland, Kuopio, Finland | Clinical Group / Task Force | |
| Eino Solje | FinnGen | University of Eastern Finland, Kuopio, Finland | Clinical Group / Task Force | |
| Hannu Kankaanranta | FinnGen | University of Gothenburg, Gothenburg, Sweden/ Seinäjoki Central | Clinical Group / Task Force | |
| Antti Mäkitie | FinnGen | University of Helsinki and Helsinki University Hospital, Helsinki, Fir | Clinical Group / Task Force | |
| Iiris Hovatta | FinnGen | University of Helsinki, Helsinki, Finland | Clinical Group / Task Force | |
| Niko Välimäki | FinnGen | University of Helsinki, Helsinki, Finland | Clinical Group / Task Force | |
| Minttu Marttila | FinnGen | University of Helsinki, Helsinki, Finland | Clinical Group / Task Force | |
| Anne Portaankorva | FinnGen | University of Helsinki, Helsinki, Finland | Clinical Group / Task Force | |
| Eija Laakkonen | FinnGen | University of Jyväskylä, Jyväskylä, Finland | Clinical Group / Task Force | |
| Heidi Silven | FinnGen | University of Oulu, Oulu, Finland | Clinical Group / Task Force | |
| Eeva Sliz | FinnGen | University of Oulu, Oulu, Finland | Clinical Group / Task Force | |

| | | | |
|---|---|---|---|
| Riikka Arffman | FinnGen | University of Oulu, Oulu, Finland | Clinical Group / Task Force |
| Susanna Savukoski | FinnGen | University of Oulu, Oulu, Finland | Clinical Group / Task Force |
| Riitta Kaarteenaho | FinnGen | University of Oulu, Oulu, Finland | Clinical Group / Task Force |
| Jaakko Tyrmi | FinnGen | University of Oulu, Oulu, Finland / University of Tampere, Tampere | Clinical Group / Task Force |
| Laura Kuusalo | FinnGen | University of Turku, Turku, Finland | Clinical Group / Task Force |
| Laura Pirilä | FinnGen | University of Turku, Turku, Finland | Clinical Group / Task Force |
| Tapio Hellman | FinnGen | University of Turku, Turku, Finland | Clinical Group / Task Force |
| Matti Vuori | FinnGen | University of Turku, Turku, Finland | Clinical Group / Task Force |
| Teemu Niiranen | FinnGen | University of Turku, Turku, Finland; Finnish Institute for Health and | Clinical Group / Task Force |
| Timo Blomster | FinnGen | Wellbeing services county of North Ostrobothnia, Oulu, Finland | Clinical Group / Task Force |
| Johanna Huhtakangas | FinnGen | Wellbeing services county of North Ostrobothnia, Oulu, Finland | Clinical Group / Task Force |
| Terttu Harju | FinnGen | Wellbeing services county of North Ostrobothnia, Oulu, Finland | Clinical Group / Task Force |
| Kaisa Tasanen | FinnGen | Wellbeing services county of North Ostrobothnia, Oulu, Finland | Clinical Group / Task Force |
| Laura Huilaja | FinnGen | Wellbeing services county of North Ostrobothnia, Oulu, Finland | Clinical Group / Task Force |
| Vuokko Anttonen | FinnGen | Wellbeing services county of North Ostrobothnia, Oulu, Finland | Clinical Group / Task Force |
| Marja Vääräsmäki | FinnGen | Wellbeing services county of North Ostrobothnia, Oulu, Finland | Clinical Group / Task Force |
| Outi Uimari | FinnGen | Wellbeing services county of North Ostrobothnia, Oulu, Finland | Clinical Group / Task Force |
| Laure Morin-Papunen | FinnGen | Wellbeing services county of North Ostrobothnia, Oulu, Finland | Clinical Group / Task Force |
| Maarit Niinimäki | FinnGen | Wellbeing services county of North Ostrobothnia, Oulu, Finland | Clinical Group / Task Force |
| Terhi Piltonen | FinnGen | Wellbeing services county of North Ostrobothnia, Oulu, Finland | Clinical Group / Task Force |
| Reetta Kälviäinen | FinnGen | Wellbeing services county of North Savo, Kuopio, Finland | Clinical Group / Task Force |
| Valtteri Julkunen | FinnGen | Wellbeing services county of North Savo, Kuopio, Finland | Clinical Group / Task Force |
| Hilkka Soininen | FinnGen | Wellbeing services county of North Savo, Kuopio, Finland | Clinical Group / Task Force |
| Mikko Kiviniemi | FinnGen | Wellbeing services county of North Savo, Kuopio, Finland | Clinical Group / Task Force |
| Oili Kaipiainen-Seppänen | FinnGen | Wellbeing services county of North Savo, Kuopio, Finland | Clinical Group / Task Force |
| Margit Pelkonen | FinnGen | Wellbeing services county of North Savo, Kuopio, Finland | Clinical Group / Task Force |
| Päivi Auvinen | FinnGen | Wellbeing services county of North Savo, Kuopio, Finland | Clinical Group / Task Force |
| Maria Siponen | FinnGen | Wellbeing services county of North Savo, Kuopio, Finland | Clinical Group / Task Force |
| Liisa Suominen | FinnGen | Wellbeing services county of North Savo, Kuopio, Finland | Clinical Group / Task Force |
| Päivi Mäntylä | FinnGen | Wellbeing services county of North Savo, Kuopio, Finland | Clinical Group / Task Force |
| Kai Kaarniranta | FinnGen | Wellbeing services county of North Savo, Kuopio, Finland; Univers | Clinical Group / Task Force |
| Jukka Peltola | FinnGen | Wellbeing Services County of Pirkanmaa, Tampere, Finland | Clinical Group / Task Force |
| Airi Jussila | FinnGen | Wellbeing Services County of Pirkanmaa, Tampere, Finland | Clinical Group / Task Force |
| Katri Kaukinen | FinnGen | Wellbeing Services County of Pirkanmaa, Tampere, Finland | Clinical Group / Task Force |
| Pia Isomäki | FinnGen | Wellbeing Services County of Pirkanmaa, Tampere, Finland | Clinical Group / Task Force |
| Jussi Hernesniemi | FinnGen | Wellbeing Services County of Pirkanmaa, Tampere, Finland | Clinical Group / Task Force |
| Annika Auranen | FinnGen | Wellbeing Services County of Pirkanmaa, Tampere, Finland | Clinical Group / Task Force |
| Hannu Uusitalo | FinnGen | Wellbeing Services County of Pirkanmaa, Tampere, Finland | Clinical Group / Task Force |
| Teea Salmi | FinnGen | Wellbeing Services County of Pirkanmaa, Tampere, Finland | Clinical Group / Task Force |
| Venla Kurra | FinnGen | Wellbeing Services County of Pirkanmaa, Tampere, Finland | Clinical Group / Task Force |
| Laura Kotaniemi-Talonen | FinnGen | Wellbeing Services County of Pirkanmaa, Tampere, Finland | Clinical Group / Task Force |
| Argyro Bizaki-Vallaskangas | FinnGen | Wellbeing Services County of Pirkanmaa, Tampere, Finland | Clinical Group / Task Force |
| Juha Rinne | FinnGen | Wellbeing Services County of Southwest Finland, Turku, Finland | Clinical Group / Task Force |
| Roosa Kallionpää | FinnGen | Wellbeing Services County of Southwest Finland, Turku, Finland | Clinical Group / Task Force |
| Markku Voutilainen | FinnGen | Wellbeing Services County of Southwest Finland, Turku, Finland | Clinical Group / Task Force |
| Antti Palomäki | FinnGen | Wellbeing Services County of Southwest Finland, Turku, Finland | Clinical Group / Task Force |
| Laura Pirilä | FinnGen | Wellbeing Services County of Southwest Finland, Turku, Finland | Clinical Group / Task Force |
| Riitta Lahesmaa | FinnGen | Wellbeing Services County of Southwest Finland, Turku, Finland | Clinical Group / Task Force |
| Kaj Metsärinne | FinnGen | Wellbeing Services County of Southwest Finland, Turku, Finland | Clinical Group / Task Force |
| Jenni Aittokallio | FinnGen | Wellbeing Services County of Southwest Finland, Turku, Finland | Clinical Group / Task Force |
| Klaus Elenius | FinnGen | Wellbeing Services County of Southwest Finland, Turku, Finland | Clinical Group / Task Force |
| Sirkku Peltonen | FinnGen | Wellbeing Services County of Southwest Finland, Turku, Finland | Clinical Group / Task Force |
| Leena Koulu | FinnGen | Wellbeing Services County of Southwest Finland, Turku, Finland | Clinical Group / Task Force |
| Ulvi Gursoy | FinnGen | Wellbeing Services County of Southwest Finland, Turku, Finland | Clinical Group / Task Force |
| Varpu Jokimaa | FinnGen | Wellbeing Services County of Southwest Finland, Turku, Finland | Clinical Group / Task Force |
| Tytti Willberg | FinnGen | Wellbeing Services County of Southwest Finland, Turku, Finland | Clinical Group / Task Force |
| Adam Ziemann | FinnGen | Abbvie, Chicago, IL, United States | Clinical Group / Task Force |
| Nizar Smaoui | FinnGen | Abbvie, Chicago, IL, United States | Clinical Group / Task Force |
| Anne Lehtonen | FinnGen | Abbvie, Chicago, IL, United States | Clinical Group / Task Force |
| Apinya Lertratanakul | FinnGen | Abbvie, Chicago, IL, United States | Clinical Group / Task Force |
| Relja Popovic | FinnGen | Abbvie, Chicago, IL, United States | Clinical Group / Task Force |
| Mengzhen Liu | FinnGen | Abbvie, Chicago, IL, United States | Clinical Group / Task Force |
| Anneke Den Hollander | FinnGen | AbbVie, Chicago, IL, United States | Clinical Group / Task Force |
| Jan Freudenberg | FinnGen | AbbVie, Chicago, IL, United States | Clinical Group / Task Force |
| Britney Milkovich | FinnGen | AbbVie, Chicago, IL, United States | Clinical Group / Task Force |
| Andrew Blumenfeld | FinnGen | AbbVie, Chicago, IL, United States | Clinical Group / Task Force |
| Tushar Kumar | FinnGen | AbbVie, Chicago, IL, United States | Clinical Group / Task Force |
| Dirk Paul | FinnGen | Astra Zeneca, Cambridge, United Kingdom | Clinical Group / Task Force |
| Bram Prins | FinnGen | Astra Zeneca, Cambridge, United Kingdom | Clinical Group / Task Force |
| Eleanor Wheeler | FinnGen | Astra Zeneca, Cambridge, United Kingdom | Clinical Group / Task Force |
| Kousik Kundu | FinnGen | Astra Zeneca, Cambridge, United Kingdom | Clinical Group / Task Force |
| Santosh Atanur | FinnGen | Astra Zeneca, Cambridge, United Kingdom | Clinical Group / Task Force |
| Andrew Lowe | FinnGen | Astra Zeneca, Cambridge, United Kingdom | Clinical Group / Task Force |
| Thomas Spargo | FinnGen | Astra Zeneca, Cambridge, United Kingdom | Clinical Group / Task Force |
| Oliver Burren | FinnGen | Astra Zeneca, Cambridge, United Kingdom | Clinical Group / Task Force |
| Margarete Fabre | FinnGen | AstraZeneca, Cambridge, United Kingdom | Clinical Group / Task Force |
| Fabio Baschiera | FinnGen | Bayer AG, Leverkusen, Germany | Clinical Group / Task Force |
| Hans van Leeuwen | FinnGen | Bayer AG, Leverkusen, Germany | Clinical Group / Task Force |
| Himanshu Manchanda | FinnGen | Bayer AG, Leverkusen, Germany | Clinical Group / Task Force |
| Karl Heilbron | FinnGen | Bayer AG, Leverkusen, Germany | Clinical Group / Task Force |
| Martin Rao | FinnGen | Bayer AG, Leverkusen, Germany | Clinical Group / Task Force |
| Nicole Schmidt | FinnGen | Bayer AG, Leverkusen, Germany | Clinical Group / Task Force |
| Samu Kurki | FinnGen | Bayer AG, Leverkusen, Germany | Clinical Group / Task Force |
| Johanna Mielke | FinnGen | Bayer AG, Leverkusen, Germany | Clinical Group / Task Force |
| Juho Immonen | FinnGen | Bayer AG, Leverkusen, Germany | Clinical Group / Task Force |
| Thomas Battram | FinnGen | Bayer AG, Leverkusen, Germany | Clinical Group / Task Force |
| Tobias Hogrebe | FinnGen | Bayer AG, Leverkusen, Germany | Clinical Group / Task Force |
| Susan Eaton | FinnGen | Biogen, Cambridge, MA, United States | Clinical Group / Task Force |
| Ketian Yu | FinnGen | Biogen, Cambridge, MA, United States | Clinical Group / Task Force |
| Stephanie Loomis | FinnGen | Biogen, Cambridge, MA, United States | Clinical Group / Task Force |
| Coro Paisan-Ruiz | FinnGen | Biogen, Cambridge, MA, United States | Clinical Group / Task Force |
| Elke Markert | FinnGen | Boehringer Ingelheim, Ingelheim am Rhein, Germany | Clinical Group / Task Force |
| Frank Li | FinnGen | Boehringer Ingelheim, Ingelheim am Rhein, Germany | Clinical Group / Task Force |
| Yao Hu | FinnGen | Boehringer Ingelheim, Ingelheim am Rhein, Germany | Clinical Group / Task Force |
| Christoph Ogris | FinnGen | Boehringer Ingelheim, Ingelheim am Rhein, Germany | Clinical Group / Task Force |

| | | | |
|---|---|---|---|
| Eric Simon | FinnGen | Boehringer Ingelheim, Ingelheim am Rhein, Germany | Clinical Group / Task Force |
| Julio Cesar Bolivar Lopez | FinnGen | Boehringer Ingelheim, Ingelheim am Rhein, Germany | Clinical Group / Task Force |
| Monika Frysz | FinnGen | Boehringer Ingelheim, Ingelheim am Rhein, Germany | Clinical Group / Task Force |
| Marla Hochfeld | FinnGen | Bristol Myers Squibb, New York, NY, United States | Clinical Group / Task Force |
| Cara Carty | FinnGen | Bristol Myers Squibb, New York, NY, United States | Clinical Group / Task Force |
| Michael Turchin | FinnGen | Bristol Myers Squibb, New York, NY, United States | Clinical Group / Task Force |
| Neelakshi Jog | FinnGen | Bristol Myers Squibb, New York, NY, United States | Clinical Group / Task Force |
| Corneliu Bodea | FinnGen | Bristol Myers Squibb, New York, NY, United States | Clinical Group / Task Force |
| Janie Shelton | FinnGen | Bristol Myers Squibb, New York, NY, United States | Clinical Group / Task Force |
| Chen Li | FinnGen | Bristol Myers Squibb, New York, NY, United States | Clinical Group / Task Force |
| Kritika Singh | FinnGen | Bristol Myers Squibb, New York, NY, United States | Clinical Group / Task Force |
| Peng Jiang | FinnGen | Bristol Myers Squibb, New York, NY, United States | Clinical Group / Task Force |
| Stephanie Loomis | FinnGen | Bristol Myers Squibb, New York, NY, United States | Clinical Group / Task Force |
| Elena Sanchez | FinnGen | Bristol Myers Squibb, New York, NY, United States | Clinical Group / Task Force |
| Lilith Moss | FinnGen | Bristol Myers Squibb, New York, NY, United States | Clinical Group / Task Force |
| Zijie Zhao | FinnGen | Bristol Myers Squibb, New York, NY, United States | Clinical Group / Task Force |
| Anna Podgornaia | FinnGen | Bristol Myers Squibb, New York, NY, United States | Clinical Group / Task Force |
| Natalie Bowers | FinnGen | Genentech, San Francisco, CA, United States | Clinical Group / Task Force |
| Edmond Teng | FinnGen | Genentech, San Francisco, CA, United States | Clinical Group / Task Force |
| Tim Lu | FinnGen | Genentech, San Francisco, CA, United States | Clinical Group / Task Force |
| Hubert Chen | FinnGen | Genentech, San Francisco, CA, United States | Clinical Group / Task Force |
| Jennifer Schutzman | FinnGen | Genentech, San Francisco, CA, United States | Clinical Group / Task Force |
| Erich Strauss | FinnGen | Genentech, San Francisco, CA, United States | Clinical Group / Task Force |
| Hao Chen | FinnGen | Genentech, San Francisco, CA, United States | Clinical Group / Task Force |
| David Choy | FinnGen | Genentech, San Francisco, CA, United States | Clinical Group / Task Force |
| Rion Pendergrass | FinnGen | Genentech, San Francisco, CA, United States | Clinical Group / Task Force |
| Brian Yaspan | FinnGen | Genentech, San Francisco, CA, United States | Clinical Group / Task Force |
| Cameron Adams | FinnGen | Genentech, San Francisco, CA, United States | Clinical Group / Task Force |
| Mark McCarthy | FinnGen | Genentech, San Francisco, CA, United States | Clinical Group / Task Force |
| Michael Rothenberg | FinnGen | Genentech, San Francisco, CA, United States | Clinical Group / Task Force |
| Rion Pendergrass | FinnGen | Genentech, San Francisco, CA, United States | Clinical Group / Task Force |
| Sergio Dellepiane | FinnGen | Genentech, San Francisco, CA, United States | Clinical Group / Task Force |
| Anubha Mahajan | FinnGen | Genentech, San Francisco, CA, United States | Clinical Group / Task Force |
| Michael Holmes | FinnGen | Genentech, San Francisco, CA, United States | Clinical Group / Task Force |
| Anubha Mahajan | FinnGen | Genentech, San Francisco, CA, United States | Clinical Group / Task Force |
| Diana Chang | FinnGen | Genentech, San Francisco, CA, United States | Clinical Group / Task Force |
| Tushar Bhangale | FinnGen | Genentech, San Francisco, CA, United States | Clinical Group / Task Force |
| Fanli Xu | FinnGen | GlaxoSmithKline, Brentford, United Kingdom | Clinical Group / Task Force |
| Laura Addis | FinnGen | GlaxoSmithKline, Brentford, United Kingdom | Clinical Group / Task Force |
| John Eicher | FinnGen | GlaxoSmithKline, Brentford, United Kingdom | Clinical Group / Task Force |
| Linda McCarthy | FinnGen | GlaxoSmithKline, Brentford, United Kingdom | Clinical Group / Task Force |
| Jorge Esparza Gordillo | FinnGen | GlaxoSmithKline, Brentford, United Kingdom | Clinical Group / Task Force |
| Joanna Betts | FinnGen | GlaxoSmithKline, Brentford, United Kingdom | Clinical Group / Task Force |
| Rajashree Mishra | FinnGen | GlaxoSmithKline, Brentford, United Kingdom | Clinical Group / Task Force |
| Audrey Chu | FinnGen | GlaxoSmithKline, Brentford, United Kingdom | Clinical Group / Task Force |
| Diptee Kulkarni | FinnGen | GlaxoSmithKline, Brentford, United Kingdom | Clinical Group / Task Force |
| Janet Kumar | FinnGen | GlaxoSmithKline, Collegeville, PA, United States | Clinical Group / Task Force |
| Charli Harlow | FinnGen | GlaxoSmithKline, Collegeville, PA, United States | Clinical Group / Task Force |
| Lea Sarow-Blat | FinnGen | GlaxoSmithKline, Collegeville, PA, United States | Clinical Group / Task Force |
| Diana L.Cousminer | FinnGen | GlaxoSmithKline, Collegeville, PA, United States | Clinical Group / Task Force |
| Jagtar Nijjar | FinnGen | GlaxoSmithKline, Collegeville, PA, United States | Clinical Group / Task Force |
| Jessica Chao | FinnGen | GlaxoSmithKline, Collegeville, PA, United States | Clinical Group / Task Force |
| Michal Magid | FinnGen | GlaxoSmithKline, Collegeville, PA, United States | Clinical Group / Task Force |
| Shashank Jariwala | FinnGen | GlaxoSmithKline, Collegeville, PA, United States | Clinical Group / Task Force |
| Chris Floyd | FinnGen | GlaxoSmithKline, Collegeville, PA, United States | Clinical Group / Task Force |
| Dan Swerdlow | FinnGen | GlaxoSmithKline, Collegeville, PA, United States | Clinical Group / Task Force |
| Erding Hu | FinnGen | GlaxoSmithKline, Collegeville, PA, United States | Clinical Group / Task Force |
| Prerak Desai | FinnGen | GlaxoSmithKline, Collegeville, PA, United States | Clinical Group / Task Force |
| Stephen Haddad | FinnGen | GlaxoSmithKline, Collegeville, PA, United States | Clinical Group / Task Force |
| Damien Croteau-Chonka | FinnGen | GlaxoSmithKline, Collegeville, PA, United States | Clinical Group / Task Force |
| Billy Fahy | FinnGen | GlaxoSmithKline, Collegeville, PA, United States | Clinical Group / Task Force |
| Paola Bronson | FinnGen | GlaxoSmithKline, Collegeville, PA, United States | Clinical Group / Task Force |
| Kirsi Auro | FinnGen | GlaxoSmithKline, Espoo, Finland | Clinical Group / Task Force |
| David Pulford | FinnGen | GlaxoSmithKline, Stevenage, United Kingdom | Clinical Group / Task Force |
| Sauli Vuoti | FinnGen | Janssen-Cilag Oy, Espoo, Finland | Clinical Group / Task Force |
| Dermot Reilly | FinnGen | Johnson & Johnson Innovative Medicine, Boston, MA, United Stat | Clinical Group / Task Force |
| Karen He | FinnGen | Johnson & Johnson Innovative Medicine, Spring House, PA, Unite | Clinical Group / Task Force |
| Ekaterina Khramtsova | FinnGen | Johnson & Johnson Innovative Medicine, Spring House, PA, Unite | Clinical Group / Task Force |
| Amy Hart | FinnGen | Johnson & Johnson Innovative Medicine, Spring House, PA, Unite | Clinical Group / Task Force |
| Meijian Guan | FinnGen | Johnson & Johnson Innovative Medicine, Spring House, PA, Unite | Clinical Group / Task Force |
| Alessandro Porello | FinnGen | Johnson & Johnson Innovative Medicine, Spring House, PA, Unite | Clinical Group / Task Force |
| P. Dunnmon | FinnGen | Johnson & Johnson Innovative Medicine, Spring House, PA, Unite | Clinical Group / Task Force |
| Sara Gale | FinnGen | Johnson & Johnson Innovative Medicine, Spring House, PA, Unite | Clinical Group / Task Force |
| Brice Keyes | FinnGen | Johnson & Johnson Innovative Medicine, Spring House, PA, Unite | Clinical Group / Task Force |
| John Kwon | FinnGen | Johnson & Johnson Innovative Medicine, Spring House, PA, Unite | Clinical Group / Task Force |
| Jonathan Sherlock | FinnGen | Johnson & Johnson Innovative Medicine, Spring House, PA, Unite | Clinical Group / Task Force |
| Matt Loza | FinnGen | Johnson & Johnson Innovative Medicine, Spring House, PA, Unite | Clinical Group / Task Force |
| Chris Whelan | FinnGen | Johnson & Johnson Innovative Medicine, Spring House, PA, Unite | Clinical Group / Task Force |
| W Galpern | FinnGen | Johnson & Johnson Innovative Medicine, Spring House, PA, Unite | Clinical Group / Task Force |
| Yanfei Zhang | FinnGen | Johnson & Johnson Innovative Medicine, Spring House, PA, Unite | Clinical Group / Task Force |
| Mona Selej | FinnGen | Johnson & Johnson Innovative Medicine, Spring House, PA, Unite | Clinical Group / Task Force |
| Abolfazl Doostparast Torshizi | FinnGen | Johnson & Johnson Innovative Medicine, Spring House, PA, Unite | Clinical Group / Task Force |
| Qingqin S Li | FinnGen | Johnson & Johnson Innovative Medicine, Titusville, NJ, United Sta | Clinical Group / Task Force |
| Sahar Mozzafari | FinnGen | Maze Therapeutics, San Francisco, CA, United States | Clinical Group / Task Force |
| Christopher Deboever | FinnGen | Maze Therapeutics, San Francisco, CA, United States | Clinical Group / Task Force |
| Jason Miller | FinnGen | Merck, Kenilworth, NJ, United States | Clinical Group / Task Force |
| Fabiana Farias | FinnGen | Merck, Kenilworth, NJ, United States | Clinical Group / Task Force |
| Andrey Loboda | FinnGen | Merck, Kenilworth, NJ, United States | Clinical Group / Task Force |
| Jorge Del-aguila | FinnGen | Merck, Kenilworth, NJ, United States | Clinical Group / Task Force |
| Elisabeth Vollmann | FinnGen | Merck, Kenilworth, NJ, United States | Clinical Group / Task Force |
| Jozsef Karman | FinnGen | Merck, Kenilworth, NJ, United States | Clinical Group / Task Force |
| Julie Fiore | FinnGen | Merck, Kenilworth, NJ, United States | Clinical Group / Task Force |
| Rajesh Kamath | FinnGen | Merck, Kenilworth, NJ, United States | Clinical Group / Task Force |
| Andrei Popescu | FinnGen | Merck, Kenilworth, NJ, United States | Clinical Group / Task Force |
| Delphine Fagegaltier | FinnGen | Merck, Kenilworth, NJ, United States | Clinical Group / Task Force |

| | | | | |
|---|---|---|---|---|
| Travis Barr | FinnGen | Merck, Kenilworth, NJ, United States | Clinical Group / Task Force | |
| Aristide Merola | FinnGen | Merck, Kenilworth, NJ, United States | Clinical Group / Task Force | |
| Oliver Freeman | FinnGen | Merck, Kenilworth, NJ, United States | Clinical Group / Task Force | |
| Simonne Longerich | FinnGen | Merck, Kenilworth, NJ, United States | Clinical Group / Task Force | |
| Enrico Ferrero | FinnGen | Novartis Institutes for BioMedical Research, Cambridge, MA, Unite | Clinical Group / Task Force | |
| Nikos Patsopoulos | FinnGen | Novartis Institutes for BioMedical Research, Cambridge, MA, Unite | Clinical Group / Task Force | |
| Nancy Finkel | FinnGen | Novartis Institutes for BioMedical Research, Cambridge, MA, Unite | Clinical Group / Task Force | |
| Sabina Pfister | FinnGen | Novartis Institutes for BioMedical Research, Cambridge, MA, Unite | Clinical Group / Task Force | |
| Shola Richards | FinnGen | Novartis Institutes for BioMedical Research, Cambridge, MA, Unite | Clinical Group / Task Force | |
| Katherine Mccauley | FinnGen | Novartis Institutes for BioMedical Research, Cambridge, MA, Unite | Clinical Group / Task Force | |
| Xiaobo Xia | FinnGen | Novartis Institutes for BioMedical Research, Cambridge, MA, Unite | Clinical Group / Task Force | |
| Mike Mendelson | FinnGen | Novartis Institutes for BioMedical Research, Cambridge, MA, Unite | Clinical Group / Task Force | |
| Majd Mouded | FinnGen | Novartis, Basel, Switzerland | Clinical Group / Task Force | |
| Debby Ngo | FinnGen | Novartis, Basel, Switzerland | Clinical Group / Task Force | |
| Kirsi Kalpala | FinnGen | Pfizer, New York, NY, United States | Clinical Group / Task Force | |
| Melissa Miller | FinnGen | Pfizer, New York, NY, United States | Clinical Group / Task Force | |
| Nan Bing | FinnGen | Pfizer, New York, NY, United States | Clinical Group / Task Force | |
| Jaakko Parkkinen | FinnGen | Pfizer, New York, NY, United States | Clinical Group / Task Force | |
| Heli Lehtonen | FinnGen | Pfizer, New York, NY, United States | Clinical Group / Task Force | |
| Stefan McDonough | FinnGen | Pfizer, New York, NY, United States | Clinical Group / Task Force | |
| Ying Wu | FinnGen | Pfizer, New York, NY, United States | Clinical Group / Task Force | |
| Erin Macdonald-Dunlop | FinnGen | Pfizer, New York, NY, United States | Clinical Group / Task Force | |
| Jessica Chung | FinnGen | Pfizer, New York, NY, United States | Clinical Group / Task Force | |
| Michael McLean | FinnGen | Pfizer, New York, NY, United States | Clinical Group / Task Force | |
| Joshua Chiou | FinnGen | Pfizer, New York, NY, United States | Clinical Group / Task Force | |
| Hye In Kim | FinnGen | Pfizer, New York, NY, United States | Clinical Group / Task Force | |
| Sivakumar Pitchumani | FinnGen | Pfizer, New York, NY, United States | Clinical Group / Task Force | |
| Sumedha Jassal | FinnGen | Pfizer, New York, NY, United States | Clinical Group / Task Force | |
| Madhurima Saxena | FinnGen | Pfizer, New York, NY, United States | Clinical Group / Task Force | |
| Catherine O'Riordan | FinnGen | Translational Sciences, Sanofi R&D, Framingham, MA, USA | Clinical Group / Task Force | |
| Samuel Lessard | FinnGen | Translational Sciences, Sanofi R&D, Framingham, MA, USA | Clinical Group / Task Force | |
| Suzanne Jacobs | FinnGen | Translational Sciences, Sanofi R&D, Framingham, MA, USA | Clinical Group / Task Force | |
| Hamid Mattoo | FinnGen | Translational Sciences, Sanofi R&D, Framingham, MA, USA | Clinical Group / Task Force | |
| David Habiel | FinnGen | Translational Sciences, Sanofi R&D, Framingham, MA, USA | Clinical Group / Task Force | |
| Guanling Huan | FinnGen | Translational Sciences, Sanofi R&D, Framingham, MA, USA | Clinical Group / Task Force | |
| Lila Kallio | FinnGen | Auria Biobank / University of Turku / Wellbeing Services County of | Biobank directors | |
| Tiina Wahlfors | FinnGen | THL Biobank / Finnish Institute for Health and Welfare (THL), Hels | Biobank directors | |
| Jukka Partanen | FinnGen | Finnish Red Cross Blood Service / Finnish Hematology Registry ar | Biobank directors | |
| Eero Punkka | FinnGen | Helsinki Biobank / Helsinki University and Hospital District of Helsi | Biobank directors | |
| Raisa Serpi | FinnGen | Northern Finland Biobank Borealis / University of Oulu / Wellbeing | Biobank directors | |
| Sanna Siltanen | FinnGen | Finnish Clinical Biobank Tampere / University of Tampere / | Biobank directors | |
| Veli-Matti Kosma | FinnGen | Biobank of Eastern Finland / University of Eastern Finland / | Biobank directors | |
| Tiina Jokela | FinnGen | Central Finland Biobank / University of Jyväskylä / Wellbeing | Biobank directors | |
| Anu Jalanko | FinnGen | Institute for Molecular Medicine Finland (FIMM), HiLIFE, University | FinnGen Teams | Administration |
| Risto Kajanne | FinnGen | Institute for Molecular Medicine Finland (FIMM), HiLIFE, University | FinnGen Teams | Administration |
| Mervi Aavikko | FinnGen | Institute for Molecular Medicine Finland (FIMM), HiLIFE, University | FinnGen Teams | Administration |
| Helen Cooper | FinnGen | Institute for Molecular Medicine Finland (FIMM), HiLIFE, University | FinnGen Teams | Administration |
| Denise Öller | FinnGen | Institute for Molecular Medicine Finland (FIMM), HiLIFE, University | FinnGen Teams | Administration |
| Tarja Laitinen | FinnGen | Institute for Molecular Medicine Finland (FIMM), HiLIFE, University | FinnGen Teams | Administration |
| Sofia Kuitunen | FinnGen | University of Helsinki, Helsinki, Finland | FinnGen Teams | Administration |
| Auli Toivola | FinnGen | Institute for Molecular Medicine Finland (FIMM), HiLIFE, University | FinnGen Teams | Sample and data logistics |
| Rodos Rodosthenous | FinnGen | Institute for Molecular Medicine Finland (FIMM), HiLIFE, University | FinnGen Teams | Sample and data logistics |
| Mitja Kurki | FinnGen | Institute for Molecular Medicine Finland (FIMM), HiLIFE, | FinnGen Teams | Analysis |
| Juha Karjalainen | FinnGen | Institute for Molecular Medicine Finland (FIMM), HiLIFE, University | FinnGen Teams | Analysis |
| Pietro Della Briotta Parolo | FinnGen | Institute for Molecular Medicine Finland (FIMM), HiLIFE, University | FinnGen Teams | Analysis |
| Arto Lehisto | FinnGen | Institute for Molecular Medicine Finland (FIMM), HiLIFE, University | FinnGen Teams | Analysis |
| Juha Mehtonen | FinnGen | Institute for Molecular Medicine Finland (FIMM), HiLIFE, University | FinnGen Teams | Analysis |
| Reza Jabal | FinnGen | Institute for Molecular Medicine Finland (FIMM), HiLIFE, | FinnGen Teams | Analysis |
| Mutaamba Maasha | FinnGen | Institute for Molecular Medicine Finland (FIMM), HiLIFE, | FinnGen Teams | Analysis |
| Sanni Ruotsalainen | FinnGen | Institute for Molecular Medicine Finland (FIMM), HiLIFE, University | FinnGen Teams | Analysis |
| Samuel Jones | FinnGen | Institute for Molecular Medicine Finland (FIMM), HiLIFE, University | FinnGen Teams | Analysis |
| Raymond Walters | FinnGen | Institute for Molecular Medicine Finland (FIMM), HiLIFE, | FinnGen Teams | Analysis |
| Paavo Häppölä | FinnGen | Institute for Molecular Medicine Finland (FIMM), HiLIFE, University | FinnGen Teams | Analysis |
| L. Elisa Lahtela | FinnGen | Institute for Molecular Medicine Finland (FIMM), HiLIFE, University | FinnGen Teams | Disease Task Forces |
| Johanna Paltta | FinnGen | University of Helsinki, Helsinki, Finland; University of Turku, | FinnGen Teams | Disease Task Forces |
| Juulia Partanen | FinnGen | Institute for Molecular Medicine Finland, HiLIFE, University of Hels | FinnGen Teams | Disease Task Forces |
| Mari Kaunisto | FinnGen | Institute for Molecular Medicine Finland (FIMM), HiLIFE, University | FinnGen Teams | Communication |
| Elina Kilpeläinen | FinnGen | Institute for Molecular Medicine Finland (FIMM), HiLIFE, University | FinnGen Teams | Sandbox & Cloud Services |
| Tianduanyi Wang | FinnGen | Institute for Molecular Medicine Finland (FIMM), HiLIFE, University | FinnGen Teams | Sandbox & Cloud Services |
| Timo P. Sipilä | FinnGen | Institute for Molecular Medicine Finland (FIMM), HiLIFE, University | FinnGen Teams | Sandbox & Cloud Services |
| Oluwaseun Alexander Dada | FinnGen | Institute for Molecular Medicine Finland (FIMM), HiLIFE, University | FinnGen Teams | Sandbox & Cloud Services |
| Awaisa Ghazal | FinnGen | Institute for Molecular Medicine Finland (FIMM), HiLIFE, University | FinnGen Teams | Sandbox & Cloud Services |
| Rigbe Weldatsadik | FinnGen | Institute for Molecular Medicine Finland (FIMM), HiLIFE, University | FinnGen Teams | Sandbox & Cloud Services |
| Jaska Uimonen | FinnGen | Institute for Molecular Medicine Finland (FIMM), HiLIFE, University | FinnGen Teams | Sandbox & Cloud Services |
| Kati Donner | FinnGen | Institute for Molecular Medicine Finland (FIMM), HiLIFE, University | FinnGen Teams | Genotyping |
| Anu Loukola | FinnGen | Helsinki Biobank / Helsinki University and Hospital District of Helsi | FinnGen Teams | Sample Collection Coordination |
| Päivi Laiho | FinnGen | THL Biobank / Finnish Institute for Health and Welfare (THL), Hels | FinnGen Teams | Sample Logistics |
| Susanna Lemmelä | FinnGen | Institute for Molecular Medicine Finland (FIMM), HiLIFE, University | FinnGen Teams | Registry Data Operations |
| Teemu Paajanen | FinnGen | THL Biobank / Finnish Institute for Health and Welfare (THL), Hels | FinnGen Teams | Registry Data Operations |
| Arto Pietilä | FinnGen | THL Biobank / Finnish Institute for Health and Welfare (THL), Hels | FinnGen Teams | Registry Data Operations |
| Aki Havulinna | FinnGen | THL Biobank / Finnish Institute for Health and Welfare (THL), Hels | FinnGen Teams | Registry Data Operations |
| Mary Pat Reeve | FinnGen | Institute for Molecular Medicine Finland (FIMM), HiLIFE, | FinnGen Teams | Phenotype team |
| Shanmukha Sampath Padmanabhuni | FinnGen | Institute for Molecular Medicine Finland (FIMM), HiLIFE, University | FinnGen Teams | Phenotype team |
| Harri Siirtola | FinnGen | University of Tampere, Tampere, Finland | FinnGen Teams | Phenotype team |
| Javier Gracia-Tabuenca | FinnGen | University of Tampere, Tampere, Finland | FinnGen Teams | Phenotype team |
| Marika Kaakinen | FinnGen | Institute for Molecular Medicine Finland (FIMM), HiLIFE, University | FinnGen Teams | Phenotype team |
| Shuang Luo | FinnGen | Institute for Molecular Medicine Finland (FIMM), HiLIFE, University | FinnGen Teams | Phenotype team |
| Vincent Llorens | FinnGen | Institute for Molecular Medicine Finland (FIMM), HiLIFE, University | FinnGen Teams | Phenotype team |
| Dawit Yohannes | FinnGen | Institute for Molecular Medicine Finland (FIMM), HiLIFE, University | FinnGen Teams | Phenotype team |
| Iina Laak | FinnGen | Institute for Molecular Medicine Finland (FIMM), HiLIFE, University | FinnGen Teams | Data protection officer |
| Mervi Ahlroth | FinnGen | Finnish Biobank Cooperative - FINBB | FinnGen Teams | FINBB - Finnish biobank cooperative |
| Johanna Mäkelä | FinnGen | Finnish Biobank Cooperative - FINBB | FinnGen Teams | FINBB - Finnish biobank cooperative |
| Pauli Wihuri | FinnGen | Finnish Biobank Cooperative - FINBB | FinnGen Teams | FINBB - Finnish biobank cooperative |
| Tom Southerington | FinnGen | Finnish Biobank Cooperative - FINBB | FinnGen Teams | FINBB - Finnish biobank cooperative |
| Meri Lähteenmäki | FinnGen | Finnish Biobank Cooperative - FINBB | FinnGen Teams | FINBB - Finnish biobank cooperative |

| | | | |
|---|---|---|---|
| Andres Metspalu | Estonian Biobank | Estonian Genome Centre, Institute of Genomics, University of Tart | **Research Team** |
| Lili Milani | Estonian Biobank | Estonian Genome Centre, Institute of Genomics, University of Tart | **Research Team** |
| Tõnu Esko | Estonian Biobank | Estonian Genome Centre, Institute of Genomics, University of Tart | **Research Team** |
| Reedik Mägi | Estonian Biobank | Estonian Genome Centre, Institute of Genomics, University of Tart | **Research Team** |
| Mait Metspalu | Estonian Biobank | Estonian Genome Centre, Institute of Genomics, University of Tart | **Research Team** |
| Mari Nelis | Estonian Biobank | Estonian Genome Centre, Institute of Genomics, University of Tart | **Research Team** |
| Georgi Hudjashov | Estonian Biobank | Estonian Genome Centre, Institute of Genomics, University of Tart | **Research Team** |
